# Antidepressant use during pregnancy and birth outcomes: analysis of electronic health data from the UK, Norway, and Sweden

**DOI:** 10.1101/2024.10.30.24316340

**Authors:** Florence Z Martin, Viktor H Ahlqvist, Paul Madley-Dowd, Michael Lundberg, Jacqueline M Cohen, Kari Furu, Dheeraj Rai, Harriet Forbes, Kayleigh Easey, Siri E Håberg, Gemma C Sharp, Cecilia Magnusson, Maria C Magnus

## Abstract

**Objectives:** To explore the association between antidepressant use during pregnancy and birth outcomes.

**Design:** Cohort study.

**Setting:** Electronic health record data.

**Participants:** 2 528 916 singleton births from the UK’s Clinical Practice Research Datalink (1996-2018), Norway’s Medical Birth Registry (2009-2020), and Sweden’s Medical Birth Register (2006-2020).

**Main outcome measures:** Stillbirth, neonatal death, pre- and post-term delivery, small and large for gestational age, and low Apgar score five minutes post-delivery.

**Results:** A total of 120 209 (4.8%) deliveries were exposed to maternal antidepressant use during pregnancy. Maternal antidepressant use during pregnancy was associated with increased odds of stillbirth (adjusted pooled OR (aOR) 1.16, 95% CI 1.05 to 1.28), preterm delivery (aOR 1.26, 95% CI 1.23 to 1.30), and Apgar score < 7 at 5 minutes (aOR 1.83, 95% CI 1.75 to 1.91). These findings persisted in the discordant sibling analysis, but with higher uncertainty. The adjusted predicted absolute risk for stillbirth was 0.34% (95% CI 0.33 to 0.35) among the unexposed and 0.40% (95% CI 0.36 to 0.44) in the antidepressant exposed. Restricting to women with depression or anxiety, the association between antidepressant exposure and stillbirth attenuated (aOR 1.07, 95% CI 0.94 to 1.21). Paternal antidepressant use was modestly associated with preterm delivery and low Apgar score. Most antidepressants were associated with preterm delivery (except paroxetine) and Apgar score (except mirtazapine and amitriptyline).

**Conclusions:** Maternal antidepressant use during pregnancy may increase the risk of stillbirth, preterm delivery, and low Apgar score, although the absolute risks remained low. Confounding by severity of indication cannot be ruled out, as the severity of symptoms was not available. The modest association between paternal antidepressant use and both preterm delivery and low Apgar score suggests that residual confounding by familial environment cannot be ruled out.

## 1 Introduction

Antidepressant use is rising globally, including during pregnancy.^1–6^ It is estimated that antidepressant prescriptions are made in as many as 8% of pregnancies in the UK,^7^ 1-2% in Norway,^8^ ^9^ and 3-4% in Sweden.^8^ The safety profile of antidepressants during pregnancy is difficult to ascertain, particularly because pregnant women are typically not included in randomised controlled trials. Observational studies are therefore the main source of evidence informing decision-making regarding antidepressant, and other, medication use during pregnancy. Confounding by indication is a pervasive problem in these studies, as studies suggest that women with untreated depression may have an increased risk of adverse birth outcomes.^10^ This is further compounded by the fact that the majority of women discontinue their antidepressant treatment during pregnancy planning, or after conception,^8^ ^11^ ^12^ and only those who have more severe symptoms continue throughout pregnancy. Those who continue their treatment into pregnancy therefore likely differ from those that do not by characteristics associated with illness severity, which might be unmeasured, and thus bias subsequent analyses.

Systematic reviews of previous observational studies support an increased risk of adverse birth outcomes, including stillbirth (17 studies, pooled odds ratio (OR) 1.19, 95% confidence interval (CI) 1.06 to 1.34), preterm delivery (13 studies, pooled OR 1.55, 95% CI 1.38 to 1.74), lower birth weight (20 studies, mean difference (MD) - 74g, 95% CI −117 to −31), and low Apgar score at 5 minutes (14 studies; standardised MD −0.33, 95% CI −0.47 to −0.20) among women who used antidepressants during pregnancy.^13^ ^14^ However, interpretability of the studies included in the above reviews may still be limited due to confounding and small sample sizes and several unanswered questions remain regarding the safety of antidepressant use during pregnancy. Firstly, the potential role of unmeasured confounding by maternal background characteristics (including socio-economic measures, underlying disorders, and genetics) needs to be clarified, as the existing studies are primarily traditionally observational, and few have leveraged approaches to infer causal relationships from such data (e.g., sibling comparisons or paternal negative control designs).^15^ Secondly, studies that have explored associations subtypes of antidepressants are limited, so it is unclear whether there might be differences in safety between different commonly used antidepressants.

We aimed to clarify whether maternal use of antidepressants during pregnancy increases the likelihood of certain birth outcomes including stillbirth, preterm delivery, and low Apgar score. The present study combined data from the UK, Norway, and Sweden to improve sample size, evaluated drug-specific effects, and leveraged a range of methods including paternal negative controls and discordant siblings to account for unobserved familial confounding.

## 2 Methods

### 2.1 Study population

We used electronic health record (EHR) data from the UK, Norway, and Sweden in the present study. Details of the data sources have been described briefly below but thoroughly elsewhere;^16–19^ data curation and harmonisation for each country are detailed in Section S1.1–S1.3. From the UK, we used the Clinical Practice Research Datalink (CPRD) GOLD, which has around 7% coverage of the UK population. CPRD GOLD contains information on prescriptions (digitalised in 1995) and diagnoses made in primary care. People who were pregnant between 1996 and 2018, had been registered with their GP for at least 12 months prior to pregnancy, and experienced a singleton delivery ≥22 weeks’ gestation were identified in the CPRD GOLD Pregnancy Register.^20^ Those who fulfilled the above criteria, as well as having linked secondary care data (∼50% of the CPRD GOLD population), were included in this study. From Norway, we included all singleton deliveries ≥22 weeks’ completed gestational weeks between 2009 and 2020 registered in the Medical Birth Registry of Norway. Dispensed prescriptions were identified from the Norwegian Prescription Database (established in 2004); diagnoses made in specialist care were identified in the Norwegian Patient Registry (established in 2008) and in primary care from the Norwegian Control and Payment of Health Reimbursement Database (KUHR, established in 2006). From Sweden, we included all deliveries ≥22 completed gestational weeks between 2006 and 2020 registered in the Medical Birth Register of Sweden. Dispensed prescriptions were identified from the Swedish Prescribed Drug Register (established in 2005) and specialist care diagnoses from the National Patient Register (established in the 1960s).

This project was approved by CPRD’s Independent Scientific Advisory Committee (ISAC) [ISAC number: 21_000362] in the UK, the Swedish Ethical Review Authority in Sweden [DNR: 2020-05516], and the Committee for Medical and Health Research Ethics of South/East Norway [2017/2546/REK sør-øst A].

### 2.2 Exposures

In the UK, prescription data were available based on the prescriptions written by general practitioners, whereas in Norway and Sweden, we used dispensation of prescription drugs from all ambulatory pharmacies (Sections S1.1.1, S1.2.4, and S1.3.3). All filled prescriptions for antidepressants were identified using the Anatomical Therapeutic Chemical (ATC) classification (N06A) that refers to antidepressants in Norway and Sweden, and the respective British National Formulary (BNF) codes (prodcodes in CPRD GOLD) in the UK (Section S1.4 and Table S1). The main exposure was any maternal antidepressant use during pregnancy proxied by prescriptions in the UK and dispensations in Norway and Sweden. We also studied use of antidepressants according to pregnancy trimester (1^st^ trimester up to gestational day 90 (12+6 weeks), 2^nd^ trimester between gestational day 91 (13 weeks) and 188 (26+6 weeks), and 3^rd^ trimester gestational day 189 (≥27 weeks) as secondary exposure. For the analysis of preterm delivery (delivery <37 completed gestational weeks), those who initiated antidepressants after gestational day 259 (i.e., no other prescriptions during pregnancy before 37 weeks’ gestation) were considered unexposed. We also further examined the nine most prescribed antidepressants by pregnant people during the study period (total number of prescriptions for each antidepressant ranked in Sweden, the largest sample). The top nine were investigated individually, with all other antidepressants combined as ‘other’: sertraline, citalopram, fluoxetine, escitalopram, venlafaxine, mirtazapine, amitriptyline, paroxetine, duloxetine, ‘other’ (Table S1), and ‘multiple’ (drug switching or concurrent prescriptions for different antidepressants).

We also retrieved corresponding data on paternal use of antidepressants during pregnancy, as a negative control exposure, in the Norwegian and Swedish data.

### 2.3 Outcomes

We examined several birth outcomes. Stillbirth was defined as fetal death upon delivery after 22–28 weeks’ gestation,^21^ the range referring to the changing definition of stillbirth over the study period in Sweden.^22^ Neonatal death was defined as death in a delivered live born during the first 28 days (0-27) of life.^23^ Due to their rarity, these outcomes were omitted from the analysis of specific antidepressants to protect patient anonymity. Preterm delivery was defined as delivery at <259 days (37 weeks’) and post-term delivery was defined as delivery ≥294 days (42 weeks’). For each week of gestation, we generated country-specific percentiles of birth weight, stratified by sex. Small for gestational age (SGA) was then defined as those with a birth weight in the lowest 10 percentiles for gestational age (<10^th^ percentile)^24^ and large for gestational age (LGA) was defined as those in the highest 10 percentiles for gestational age (>90^th^ percentile)^25^ as per our country-specific curves. In the Norway and Sweden, Apgar score is reported in the medical birth registries at 1, 5, and 10 minutes.^17^ ^23^ Apgar score at 5 minutes was chosen here, due to its clinical correlation with mortality and cerebral palsy risk.^26^ It was further binarized into <7 and ≥7, as scores 7-10 are considered reassuring.^26^ Information on neonatal death and Apgar score was not available for UK. In Sweden, there was no paternal linkage with stillborn babies.

### 2.4 Covariates

Given the varying degrees of data availability in each country, we utilised a country optimised adjustment approach^27^ where covariates common to all countries were included in all models and additional covariates were added in a country-specific manner.

Common covariates included maternal age at delivery (<20, 20-24, 25-29, 30-34, 35-39, 40-44 and ≥45), early-pregnancy body mass index (BMI; underweight <18kg/m^2^, ‘normal’ weight 18-24.9 kg/m^2^, overweight 25-29.9 kg/m^2^, and obese ≥30 kg/m^2^), parity (categorical: 0, 1, 2, 3, and ≥4), previous stillbirth (binary), anti-seizure medication and antipsychotic use in the 12 months prior to pregnancy (binary), smoking anytime during pregnancy (binary), and maternal depression or anxiety diagnosis prior to the start of pregnancy (binary, ≥1 code any time during pre-pregnancy follow-up). Administrative codes were used to ascertain the presence of a potential indication for antidepressant prescription (evidence of a depression or anxiety diagnosis: ICPC2 codes from primary care/ICD-10 codes from secondary care in Norway, Read codes from primary care/ICD-10 codes from secondary care in the UK, and ICD-9/ICD-10 codes from secondary care in Sweden), and to define prescription/dispensation of anti-seizure and antipsychotic medications (BNF codes in the UK and ATC codes in Norway and Sweden). All codes are summarised as codelists in https://github.com/flozoemartin/codelists.

We also retrieved different proxy measures of socioeconomic position (SEP). This included maternal educational attainment (categorical: compulsory or less, secondary, post-secondary, post-graduate, and missing) and country of birth (binarized into foreign-born or not) in Norway and Sweden, household disposable income according to quintiles in Sweden, and practice-level Index of Multiple Deprivation (IMD) quintile and ethnicity (White, South Asian, Black, Other, Mixed)^28^ in the UK.

We used a complete records approach and used a country-optimised adjustment approach to account for confounding, leveraging the covariates available in each country specifically.^27^ Covariates that had >5% missing were dropped from the primary analysis adjustment set in each country then added back in in sensitivity analysis. The confounders included in the primary models in each country and covariates dropped due to missing data are summarised in Table S2.

### 2.5 Analysis

We used logistic regression to estimate adjusted ORs and absolute (standardized) risks of the outcomes separately in each country. We replicated these analyses for trimester-specific antidepressant use and mono- and polytherapy of the top prescribed antidepressants. These were then combined using Mantel-Haenszel weighted meta-analysis with fixed effects. Standardised mean differences were obtained from multivariable linear regression models for continuous outcomes: gestational age (week) and birthweight (grams).

To control for shared genetic and household factors, as well as stable maternal factors between pregnancies, we used a discordant sibling design implemented by conditional logistic regression. Failing to replicate an association within families may suggest that an observed association in the general population could be explained by maternal and household confounders which remain stable between pregnancies.^29^ We also evaluated the association with paternal use of antidepressants while the mother was pregnant, to clarify the potential the role of residual (household) confounding.^30^ We additionally performed mutual adjustment for maternal antidepressant use in sensitivity analysis to limit bias by assortative mating.^31^ These analyses were possible in Norway and Sweden only.

We conducted several sensitivity analyses to evaluate the robustness of our findings. Given that we dropped covariates from the primary adjustment set if they had >5% missing data (Table S2), we repeated the primary maternal analysis adjusting for all covariates (irrespective of the proportion of missing data) in sensitivity analyses and tested for potential bias in the complete records analysis as per Hughes *et al*.^32^ To investigate the approach that we used to manage confounding by indication, we evaluated the associations with maternal antidepressant use stratified on indication (depression or anxiety), rather than adjusting for it.^33^ Given that infant morbidity and mortality changes as gestational age at delivery drops, we stratified preterm delivery according to moderate-to-late preterm (≥32 and <37 weeks’ gestation), very preterm (≥28 and <32 weeks’), and extremely preterm (<28 weeks’). To explore the role of labour induction, we also investigated the risk of preterm and post-term delivery restricted to those who delivered spontaneously. We re-ran the analysis for SGA and LGA using definitions as per the INTERGROWTH-21 standard ^34^ and the new Swedish standard.^35^

Several more data management decisions had been made when cleaning the UK data, so a number of sensitivity analyses were performed to test these, including using alternative sources of information to assign pre- and post-term delivery and changing the threshold of post-term delivery to reflect the imputation approach leveraged by the authors of the Pregnancy Register algorithm.^20^ These analyses are detailed in the supplement.

All data cleaning, analyses, and data visualisations were performed in Stata 17 and all scripts are available on GitHub (https://github.com/flozoemartin/Birth-outcomes).

### 2.6 Patient and public involvement

Patients were not consulted during the design, conduct, or reporting of the present study due to time limitations within the funded PhD programme that the work was conducted during. Results will be disseminated to the public via this publication, social media, and clinical practice.

## 3 Results

The eligible study population contained 352 361 singleton deliveries from the UK, 679 511 from Norway, and 1 497 044 from Sweden (Figure S1). A total of 120 209 (4.8%) pregnancies were exposed to antidepressants; 6.2% of the UK’s deliveries, 2.5% of Norway’s, and 5.4% of Sweden’s.

Maternal background characteristics were broadly similar across the three countries (Table 1). An important difference between the countries was that prevalence of registered depression and anxiety diagnoses was lower in Sweden, which reflects the lack of information on diagnoses made in primary care data in Sweden (Table 1). Differences in socioeconomic measures observed between countries are shown in Table S3, where in all three countries, proxies of lower SEP (lower educational attainment or higher deprivation quintile) were associated with antidepressant use during pregnancy.

**Table 1.**
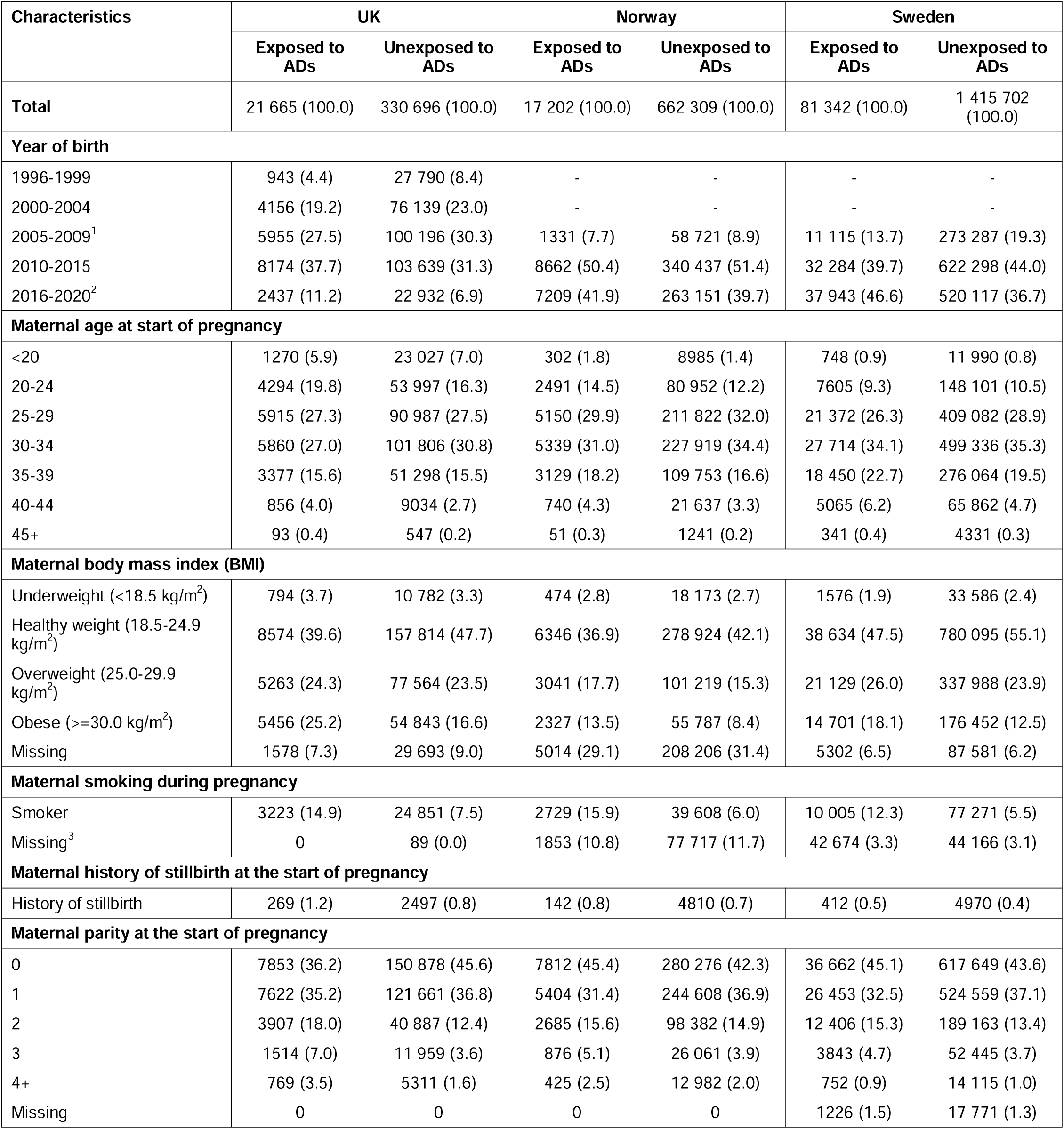
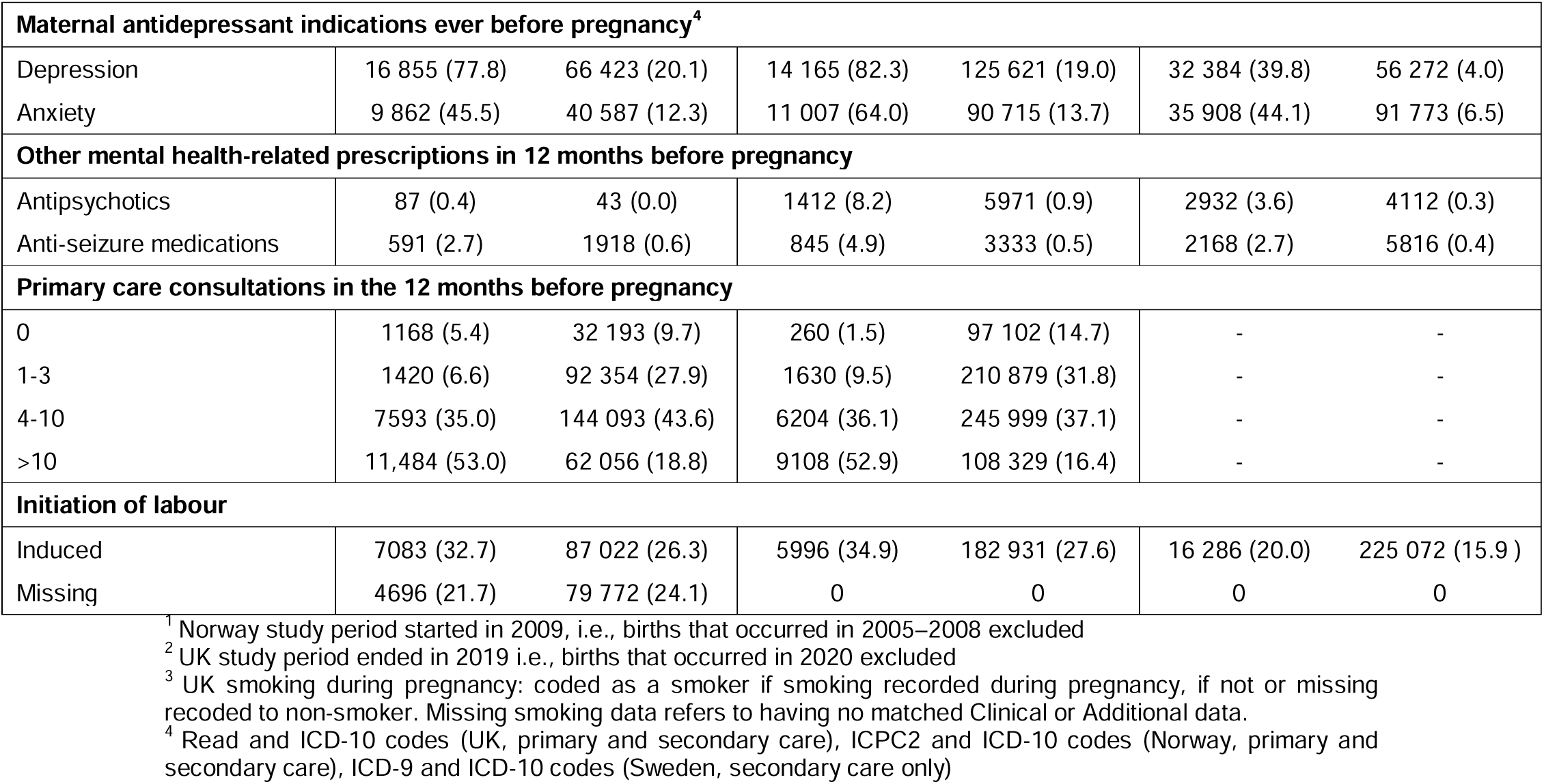
Maternal characteristics in eligible populations from each country, stratified by antidepressant exposure status during pregnancy.

| Characteristics | UK |  | Norway |  | Sweden |  |
| --- | --- | --- | --- | --- | --- | --- |
|  | Exposed to<br>ADs | Unexposed to<br>ADs | Exposed to<br>ADs | Unexposed to<br>ADs | Exposed to<br>ADs | Unexposed to<br>ADs |
| **Total** | 21 665 (100.0) | 330 696 (100.0) | 17 202 (100.0) | 662 309 (100.0) | 81 342 (100.0) | 1 415 702<br>(100.0) |
| **Year of birth** |  |  |  |  |  |  |
| 1996-1999 | 943 (4.4) | 27 790 (8.4) | - | - | - | - |
| 2000-2004 | 4156 (19.2) | 76 139 (23.0) | - | - | - | - |
| 2005-2009<sup>1</sup> | 5955 (27.5) | 100 196 (30.3) | 1331 (7.7) | 58 721 (8.9) | 11 115 (13.7) | 273 287 (19.3) |
| 2010-2015 | 8174 (37.7) | 103 639 (31.3) | 8662 (50.4) | 340 437 (51.4) | 32 284 (39.7) | 622 298 (44.0) |
| 2016-2020<sup>2</sup> | 2437 (11.2) | 22 932 (6.9) | 7209 (41.9) | 263 151 (39.7) | 37 943 (46.6) | 520 117 (36.7) |
| **Maternal age at start of pregnancy** |  |  |  |  |  |  |
| <20 | 1270 (5.9) | 23 027 (7.0) | 302 (1.8) | 8985 (1.4) | 748 (0.9) | 11 990 (0.8) |
| 20-24 | 4294 (19.8) | 53 997 (16.3) | 2491 (14.5) | 80 952 (12.2) | 7605 (9.3) | 148 101 (10.5) |
| 25-29 | 5915 (27.3) | 90 987 (27.5) | 5150 (29.9) | 211 822 (32.0) | 21 372 (26.3) | 409 082 (28.9) |
| 30-34 | 5860 (27.0) | 101 806 (30.8) | 5339 (31.0) | 227 919 (34.4) | 27 714 (34.1) | 499 336 (35.3) |
| 35-39 | 3377 (15.6) | 51 298 (15.5) | 3129 (18.2) | 109 753 (16.6) | 18 450 (22.7) | 276 064 (19.5) |
| 40-44 | 856 (4.0) | 9034 (2.7) | 740 (4.3) | 21 637 (3.3) | 5065 (6.2) | 65 862 (4.7) |
| 45+ | 93 (0.4) | 547 (0.2) | 51 (0.3) | 1241 (0.2) | 341 (0.4) | 4331 (0.3) |
| **Maternal body mass index (BMI)** |  |  |  |  |  |  |
| Underweight (<18.5 kg/m<sup>2</sup>) | 794 (3.7) | 10 782 (3.3) | 474 (2.8) | 18 173 (2.7) | 1576 (1.9) | 33 586 (2.4) |
| Healthy weight (18.5-24.9 kg/m<sup>2</sup>) | 8574 (39.6) | 157 814 (47.7) | 6346 (36.9) | 278 924 (42.1) | 38 634 (47.5) | 780 095 (55.1) |
| Overweight (25.0-29.9 kg/m<sup>2</sup>) | 5263 (24.3) | 77 564 (23.5) | 3041 (17.7) | 101 219 (15.3) | 21 129 (26.0) | 337 988 (23.9) |
| Obese (≥30.0 kg/m<sup>2</sup>) | 5456 (25.2) | 54 843 (16.6) | 2327 (13.5) | 55 787 (8.4) | 14 701 (18.1) | 176 452 (12.5) |
| Missing | 1578 (7.3) | 29 693 (9.0) | 5014 (29.1) | 208 206 (31.4) | 5302 (6.5) | 87 581 (6.2) |
| **Maternal smoking during pregnancy** |  |  |  |  |  |  |
| Smoker | 3223 (14.9) | 24 851 (7.5) | 2729 (15.9) | 39 608 (6.0) | 10 005 (12.3) | 77 271 (5.5) |
| Missing<sup>3</sup> | 0 | 89 (0.0) | 1853 (10.8) | 77 717 (11.7) | 42 674 (3.3) | 44 166 (3.1) |
| **Maternal history of stillbirth at the start of pregnancy** |  |  |  |  |  |  |
| History of stillbirth | 269 (1.2) | 2497 (0.8) | 142 (0.8) | 4810 (0.7) | 412 (0.5) | 4970 (0.4) |
| **Maternal parity at the start of pregnancy** |  |  |  |  |  |  |
| 0 | 7853 (36.2) | 150 878 (45.6) | 7812 (45.4) | 280 276 (42.3) | 36 662 (45.1) | 617 649 (43.6) |
| 1 | 7622 (35.2) | 121 661 (36.8) | 5404 (31.4) | 244 608 (36.9) | 26 453 (32.5) | 524 559 (37.1) |
| 2 | 3907 (18.0) | 40 887 (12.4) | 2685 (15.6) | 98 382 (14.9) | 12 406 (15.3) | 189 163 (13.4) |
| 3 | 1514 (7.0) | 11 959 (3.6) | 876 (5.1) | 26 061 (3.9) | 3843 (4.7) | 52 445 (3.7) |
| 4+ | 769 (3.5) | 5311 (1.6) | 425 (2.5) | 12 982 (2.0) | 752 (0.9) | 14 115 (1.0) |
| Missing | 0 | 0 | 0 | 0 | 1226 (1.5) | 17 771 (1.3) |

| **Maternal antidepressant indications ever before pregnancy<sup>4</sup>** |  |  |  |  |  |  |
| --- | --- | --- | --- | --- | --- | --- |
| Depression | 16 855 (77.8) | 66 423 (20.1) | 14 165 (82.3) | 125 621 (19.0) | 32 384 (39.8) | 56 272 (4.0) |
| Anxiety | 9 862 (45.5) | 40 587 (12.3) | 11 007 (64.0) | 90 715 (13.7) | 35 908 (44.1) | 91 773 (6.5) |
| **Other mental health-related prescriptions in 12 months before pregnancy** |  |  |  |  |  |  |
| Antipsychotics | 87 (0.4) | 43 (0.0) | 1412 (8.2) | 5971 (0.9) | 2932 (3.6) | 4112 (0.3) |
| Anti-seizure medications | 591 (2.7) | 1918 (0.6) | 845 (4.9) | 3333 (0.5) | 2168 (2.7) | 5816 (0.4) |
| **Primary care consultations in the 12 months before pregnancy** |  |  |  |  |  |  |
| 0 | 1168 (5.4) | 32 193 (9.7) | 260 (1.5) | 97 102 (14.7) | - | - |
| 1-3 | 1420 (6.6) | 92 354 (27.9) | 1630 (9.5) | 210 879 (31.8) | - | - |
| 4-10 | 7593 (35.0) | 144 093 (43.6) | 6204 (36.1) | 245 999 (37.1) | - | - |
| >10 | 11,484 (53.0) | 62 056 (18.8) | 9108 (52.9) | 108 329 (16.4) | - | - |
| **Initiation of labour** |  |  |  |  |  |  |
| Induced | 7083 (32.7) | 87 022 (26.3) | 5996 (34.9) | 182 931 (27.6) | 16 286 (20.0) | 225 072 (15.9 ) |
| Missing | 4696 (21.7) | 79 772 (24.1) | 0 | 0 | 0 | 0 |

<sup>1</sup> Norway study period started in 2009, i.e., births that occurred in 2005–2008 excluded
<sup>2</sup> UK study period ended in 2019 i.e., births that occurred in 2020 excluded
<sup>3</sup> UK smoking during pregnancy: coded as a smoker if smoking recorded during pregnancy, if not or missing recoded to non-smoker. Missing smoking data refers to having no matched Clinical or Additional data.
<sup>4</sup> Read and ICD-10 codes (UK, primary and secondary care), ICPC2 and ICD-10 codes (Norway, primary and secondary care), ICD-9 and ICD-10 codes (Sweden, secondary care only)

Overall, the rates of most outcomes were similar between the three countries. Among unexposed pregnancies, 0.3-0.5% ended in stillbirth, 4.7-6.6% were born preterm and 4.5-6.7% were born post-term. Among the exposed, 0.4-0.6% ended in stillbirth, 7.3-8.6% were preterm, and 3.1-4.7% were post-term (Table S4 and Table S5).

### 3.1 Maternal analysis

In the fixed effects meta-analysis of maternal antidepressant use during pregnancy, we observed an increased risk of stillbirth (adjusted OR (aOR) 1.16, 95% CI 1.05 to 1.28). The adjusted absolute risk of stillbirth was 0.34% (95% CI 0.33 to 0.35) in the unexposed and 0.40% (95% CI 0.36 to 0.44) in the antidepressant exposed. We also observed an increased risk of preterm delivery (aOR 1.26, 95% CI 1.23 to 1.30), SGA (aOR 1.04, 95% CI 1.02 to 1.07), and Apgar score <7 at 5 minutes (aOR 1.83, 95% CI 1.75 to 1.91), as well as a decreased risk of post-term delivery (aOR 0.75, 95% CI 0.72 to 0.77) and no notable difference in the risk of LGA nor neonatal death (Figure 1). Effect estimates from each country contributing to each analysis can be found in Figure S2.

**Figure 1.**
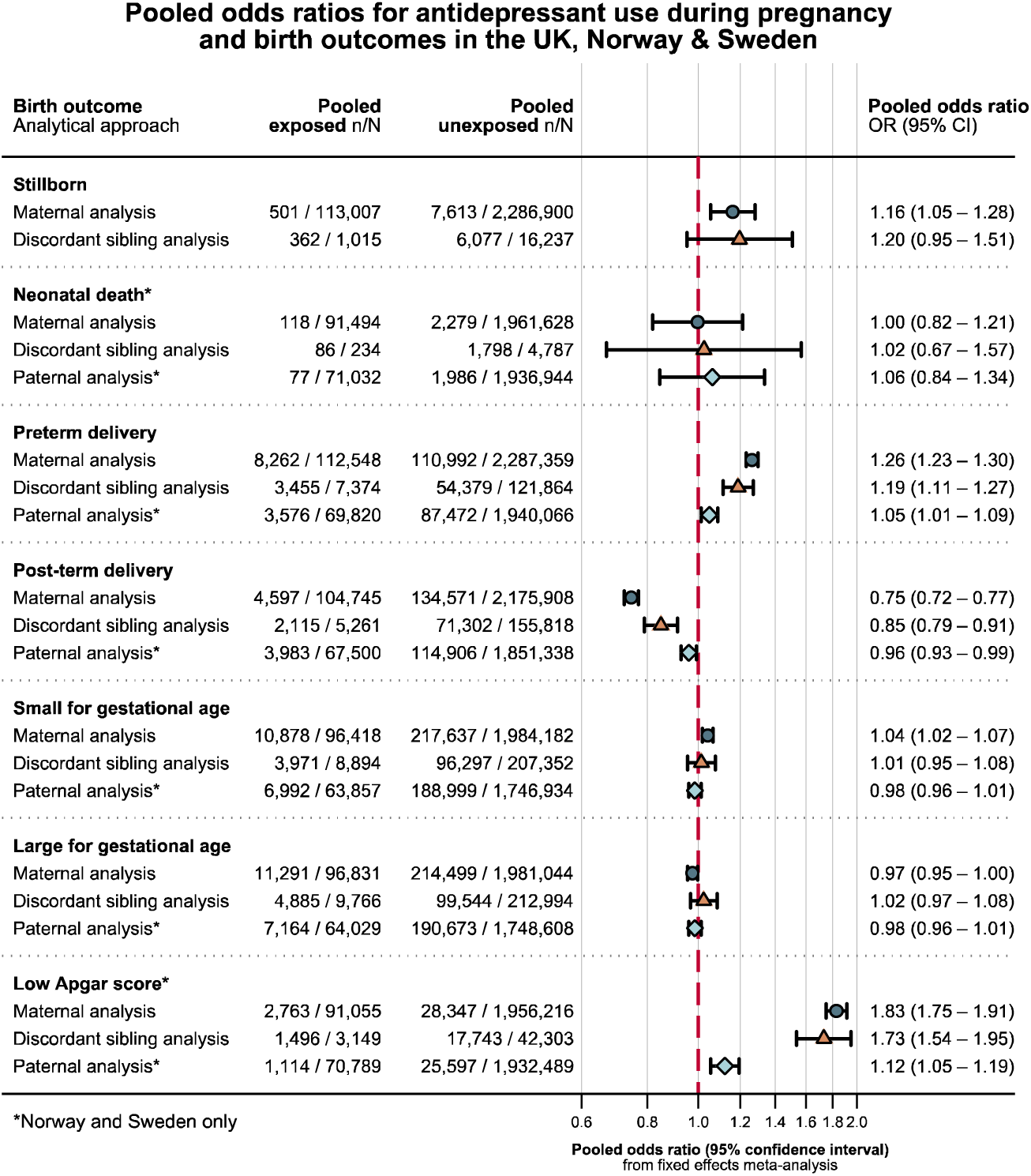
Primary analysis of maternal antidepressant use during pregnancy^1^, maternal antidepressant use among discordant siblings^2^, and paternal antidepressant use during pregnancy^3^ as a negative control.

The trimester-specific analysis indicated the risk of low Apgar score increased with trimester of exposure and risk of post-term decreased (Figure S3). Most antidepressants were associated with preterm delivery and Apgar score <5 minutes (Figure 2, Figure S4, Figure S6). The use of citalopram, fluoxetine, mirtazapine, or paroxetine during pregnancy was also associated with a higher risk of SGA (Figure 2, Figure S5). The increased risk translated to modest differences in absolute risk adjusted for confounders across all medications (Figure 3) and adjusted mean differences in gestational week and weight at delivery (Table S6 and Table S7).

**Figure 2.**
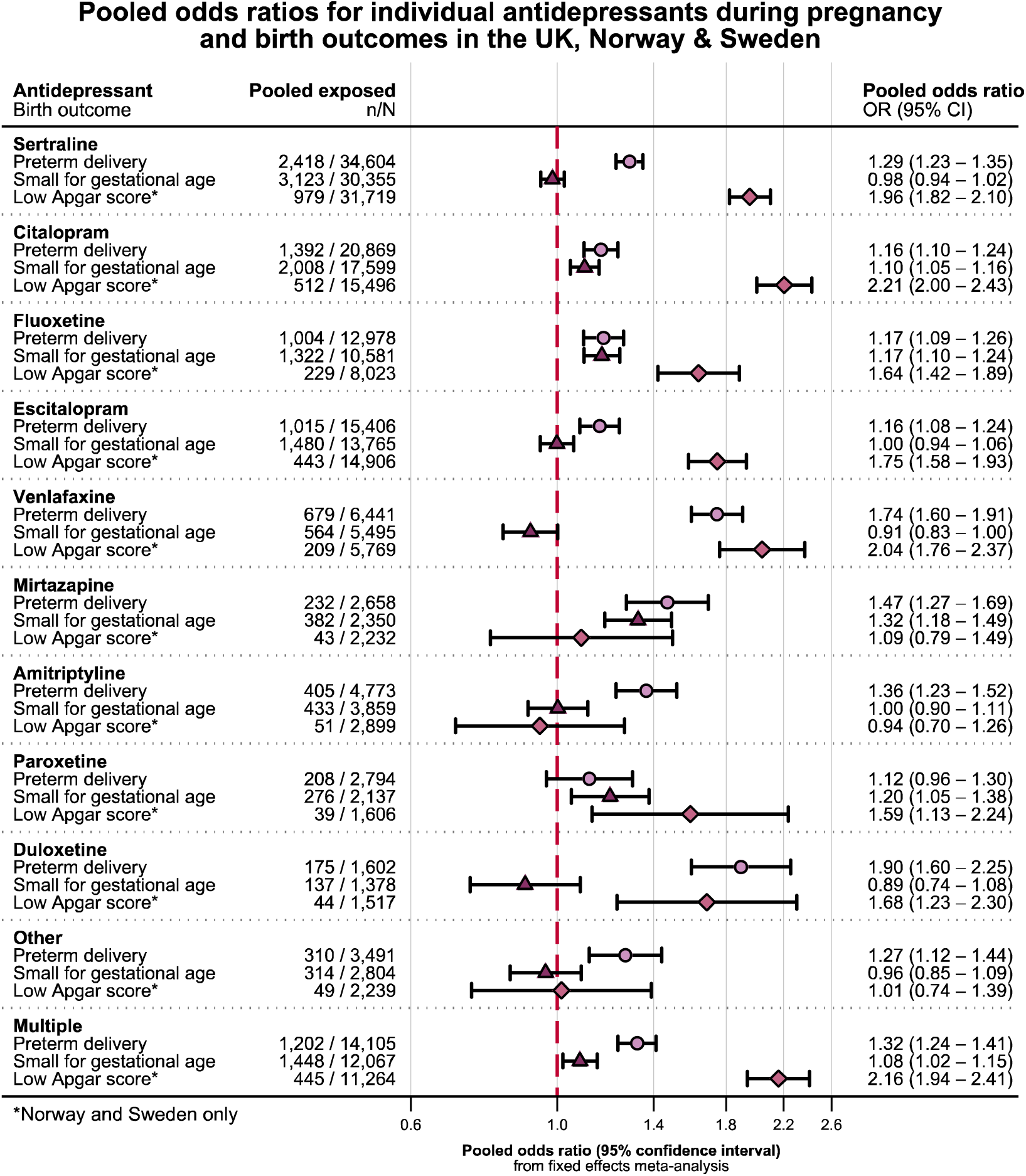
Drug-specific analyses^4^ for preterm delivery, small for gestational age, and Apgar score < 7 at 5 minutes after delivery.

**Figure 3.**
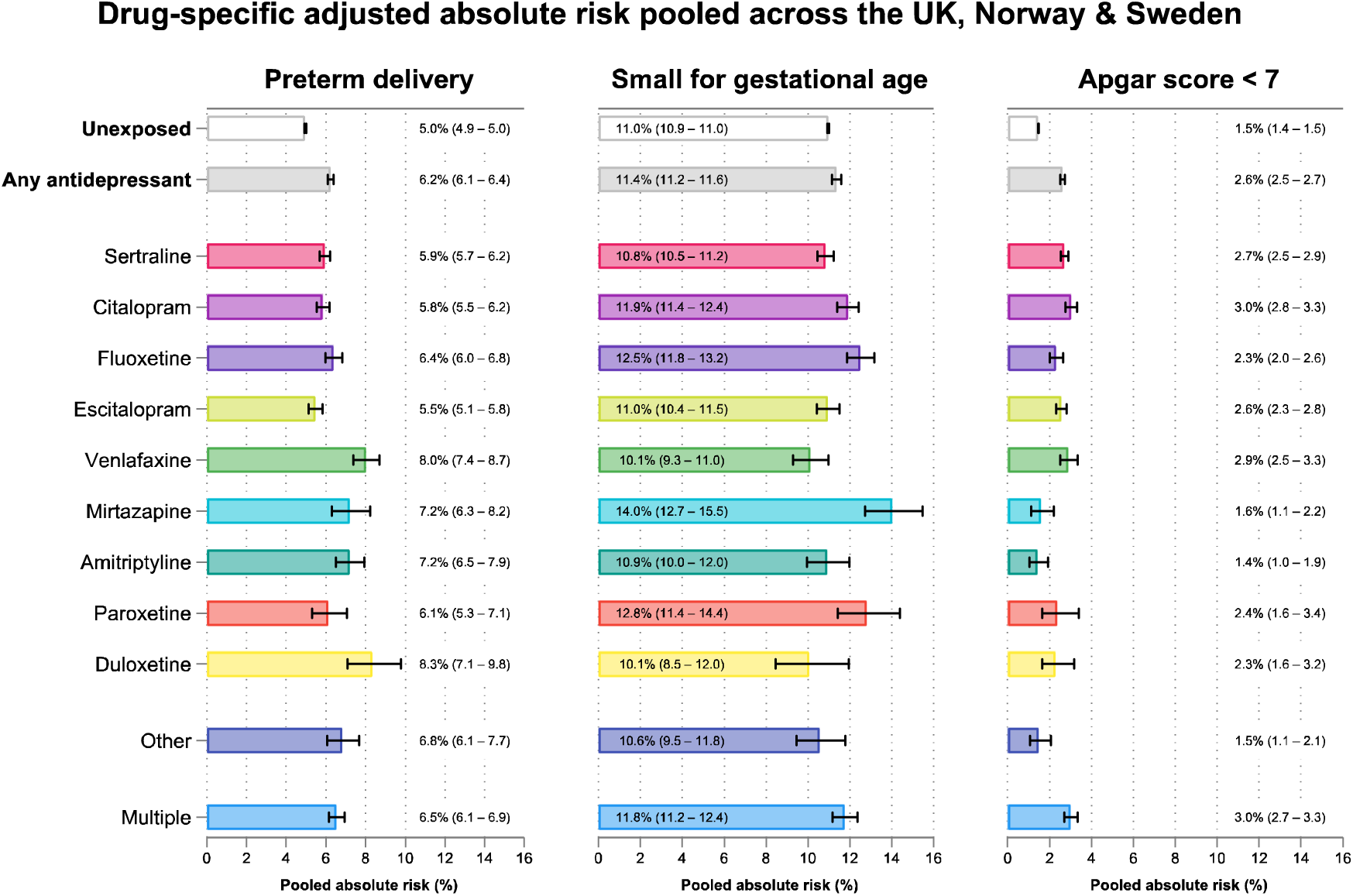
Pooled absolute risk adjusted for confounders^5^ of preterm delivery, SGA, and low Apgar score for each antidepressant medication.

### 3.2 Discordant sibling analysis

The number of exposure and outcome discordant sibling pairs available for the different analyses ranged from 419 to 2743 in the UK, 108 to 2155 in Norway, and 273 to 9794 in Sweden (Table S8). The sibling analyses yielded similar results to the primary analysis, albeit with wider confidence intervals (Figure 1, Figure S7).

### 3.3 Paternal negative control analysis

Information on paternal use of antidepressants was available for 2 127 142 deliveries in Norway and Sweden. Fathers used antidepressants while the mother was pregnant in 76 080 (3.6%) of the deliveries included in the sample. Paternal background characteristics according to use of antidepressants in Norway and Sweden (Table S9). The associations with paternal use of antidepressants were null for most outcomes (Figure 1, Figure S8). The upper CI remained below one for post-term delivery and the lower CI remained above one for preterm delivery and low Apgar score. Further adjustment for maternal use of antidepressants did not change our conclusions for paternal antidepressant use (Figure S9).

### 3.4 Sensitivity analyses

We observed similar associations when adding ethnicity and BMI (UK) and smoking (Norway) to the primary analysis model (Figure S10). When restricting the populations to those with evidence of depression or anxiety prior to pregnancy, we observe similar results to the primary analysis, although an attenuation to the null for stillbirth was seen in Sweden (Figure S11). Antidepressant use was associated with increased risk of ‘moderate-to-late’ preterm delivery, but not ‘very’ or ‘extremely’ preterm delivery (Figure S12), and an increased risk of both spontaneous and induced preterm delivery (Figure S13). When restricting to spontaneous deliveries in the evaluation of the risk of post-term birth, the reduced risk observed following maternal antidepressant use in the main analysis slightly attenuated (Figure S14). Using the INTERGROWTH-21 and new Swedish standards (Table S1), we observed similar results to the primary analysis for SGA and LGA (Table S10). The UK-specific sensitivity analyses are summarised in Tables S11–S13. To note, using different methods to define post-term birth, including gestational age at birth reported in secondary care or codes that denoted the delivery as ‘late’, we observed findings in the UK that reflected the suggested protective association observed in Norway and Sweden (Table S12).

## 4 Discussion

### 4.1 Statement of principal findings

In this large study including data from UK, Norway, and Sweden, we found that women who used antidepressants during pregnancy had an increased risk of stillbirth, preterm delivery, and low Apgar score at five minutes post-delivery. The lower risk of post-term delivery among women who had used antidepressants in the main analysis was not completely explained by induction of labour and warrants further investigation.

In response to calls from previous studies, we investigated trimester-specific exposure.^36^ As trimester of medication use increases, antidepressants appear more protective for post-term delivery and LGA and increase the risk of preterm delivery, SGA, and low Apgar score. However, the confidence intervals for the trimester-specific exposure were mostly overlapping, and differences may be attributable to confounding by severity of indication (whereby those who are more unwell are more likely to continue their regimen throughout more of their pregnancy) and differences in obstetric care between those with and without a ‘complicated’ pregnancy.

Each of the studied antidepressants appeared to be associated with an increased risk of preterm birth (although the estimate for paroxetine was imprecise), and most were associated with a low Apgar score (except mirtazapine, amitriptyline, and ‘other’ antidepressants). The associations between individual drugs and SGA were mixed, where citalopram, fluoxetine, mirtazapine, paroxetine, and ‘multiple’ were associated with an increased risk, whereas sertraline, escitalopram, duloxetine, venlafaxine, and ‘other’ were not. Confounding by severity of indication can be inherent to these analyses, considering that SSRIs tend to be first line treatments, while SNRIs, monoamine oxidase inhibitors, and atypical antidepressants are only prescribed if SSRIs are not effective. We observed a stronger association for non-SSRI medications than SSRIs in the analysis of preterm delivery, which could be compatible with such confounding by severity of indication. The confounding in these analyses may, however, be tempered by the combination of differing prescribing patterns between countries, for example escitalopram is the most prevalently prescribed antidepressant in Norway, but more rarely prescribed in the UK and Sweden.

### 4.2 Strengths and weaknesses

The present study has several strengths. It leverages data from three countries, two of which include all births in the respective countries, linked with dispensation data. Due to the size of these data when pooled in meta-analysis, and within-family linkage capabilities (i.e., linking siblings with mothers), we were able to use multiple family-based methods, namely paternal negative exposure control analyses (in Norway and Sweden) and discordant sibling analyses. The study also has some limitations. We used a combination of prescriptions written (UK) and prescriptions dispensed (Norway and Sweden) to assign exposure status. There is likely to be some exposure misclassification if the medication wasn’t taken, especially in the UK where we do not know if the prescription was dispensed,^37^ however it is up for debate how impactful non-differential exposure misclassification is.^38^ Furthermore, if Apgar score assignment differed systematically in light of assessor knowledge of maternal antidepressant use (e.g., assessors paying closer attention to children exposed to medications), this could introduce differential outcome misclassification. The definition of the exposure as “any during pregnancy” does not reflect real-world prescribing of antidepressants^39^ and does not take into consideration things like discontinuation, a very common pattern of prescribing during pregnancy.^12^ We did however attempt to mitigate this by performing a trimester-specific analysis.

The findings from the sibling analysis mostly reflect the findings from the primary maternal analysis. However, these are still sensitive to all confounders that are not shared across the pregnancies,^29^ such as any new-onset disease. We found mostly null results in the paternal negative exposure control analysis, leading us to be cautiously reassured that our shared environment confounding has been adequately dealt with in the primary maternal analysis. However, for preterm delivery and Apgar score, we cannot rule out the possibility of residual confounding based on the negative control analysis. One of the main confounding barriers that affect drug safety analysis in pregnancy is confounding by severity of indication. Firstly, we used evidence of a diagnosis of depression or anxiety to proxy potential antidepressant indication, but we don’t know from EHR data that any one diagnosis refers to the indication to a prescribed medication. Depression and anxiety are diseases with variable clinical trajectories,^40^ with periods of remission and relapse, thus it is likely that exposure to antidepressants is in part proxying periods of “active” illness. To note, indication was obtained differently across countries: unlike the UK and Norway, primary care data were not available in Sweden, meaning that indication was obtained only for those who had been hospitalised or sought specialist care for depression or anxiety. This likely resulted in only those who were more unwell being flagged as having depression or anxiety in Sweden.

### 4.3 Comparison to the literature

Our findings are in line with existing evidence from meta-analyses indicating a greater risk of stillbirth, preterm birth, and low Apgar score among women who have used antidepressants during pregnancy.^13^ ^14^ ^41^ Our finding for stillbirth aligns closely with the systematic review findings from Martin *et al*. (aOR 1.16 *v* summary effect estimate 1.19^13^). However, unlike our study, some individual studies of stillbirth have concluded that antidepressants are not associated with an increase in risk of stillbirth or neonatal death.^42^ ^43^ Specifically, Stephansson *et al.* showed that the crude odds ratio >1 for stillbirth and neonatal death attenuated to the null when restricting to those with a previous psychiatric hospitalization.^43^ Finally, we observed little association with size for gestational age, in line with the findings from Sujan *et al.*^36^ but in conflict with other papers that did find a modest association with SGA.^44^ ^45^ Our findings are also consistent with previous studies that have reported associations between SSRIs and low Apgar score,^14^ ^46^ and show evidence of an association with venlafaxine and duloxetine (SNRIs) with low Apgar score, but not amitriptyline (TCA) or mirtazapine (atypical). Antidepressant use, particularly SSRIs, during pregnancy has been previously associated with delayed neonatal adaptation,^47^ a symptom of which is low Apgar score. We also show that antidepressant use is associated with preterm delivery, which itself is associated with low Apgar score.^48^

The consistent results from the sibling analyses indicate that our findings are unlikely to be explained by unmeasured confounding by maternal characteristics which remain stable between deliveries. However, confounding by indication cannot be ruled out in these analyses, as symptoms of depression vary over time. The fact that we did not observe an increased risk of stillbirth, neonatal death, or size for gestational age with paternal use of antidepressants lends further support to the notion that unmeasured confounding may not explain the primary maternal results.

A biological effect of maternal antidepressant use during pregnancy on these birth outcomes is plausible. Serotonin plays an important role in fetal development^49^ and circulating (free) serotonin is higher in those taking antidepressants, particularly those that block serotonin reuptake, like SSRIs.^50^ Animal studies have shown serotonin exposure can lead to placental inflammation,^51^ which is associated with both stillbirth and preterm delivery.^52^ However, maternal stress also interacts with the serotonergic system,^53^ making it even more difficult to disentangle confounding by indication.

### 4.4 Clinical Implications

The current advice for antidepressant use during pregnancy varies from country to country and is tailored to individual patients. The findings from this study suggest that antidepressants are associated with a modest increased risk of stillbirth, preterm delivery, and low Apgar score. Venlafaxine and duloxetine were more strongly associated with preterm delivery and mirtazapine and paroxetine for SGA. However, we would urge caution in interpreting these findings as causal effects that could be used to inform clinical decision making, given the challenge of extricating the effect of the medication from the total effect of the medication and the indication (e.g., antidepressants to treat depression). In addition, the absolute risk of each outcome for pregnancies exposed to antidepressants remain small as compared to unexposed pregnancies. For example, 5.0% of unexposed pregnancies compared to 6.2% of exposed pregnancies were born preterm, when taking account for confounders. Any decisions regarding antidepressant use during pregnancy should weigh the risk resulting from untreated psychiatric conditions where antidepressants may be effective, against the increased risk of adverse outcomes found in this study.

### 4.5 Future work

Data sources that consider disease severity are crucial to better understand the fetal safety of antidepressants that is agnostic to indication. Once we are able to better account for indication in pregnancy pharmacoepidemiology, we will be able to interpret causal inference methods more robustly.

### 4.6 Conclusions

Maternal antidepressant use during pregnancy is associated with an increased risk of stillbirth, preterm delivery, and low Apgar score, although the absolute risk for these outcomes remains low. Despite using multiple approaches to infer causal effects from observational data, residual confounding by severity of indication, or household-level factors, is difficult to rule out. Our study still emphasizes the importance of considering fetal safety when prescribing antidepressants during pregnancy.

## Supporting information

Section S, Table S, Figure S

## Data Availability

Access to CPRD data, including UK Primary Care Data, and linked data such as Hospital Episode Statistics, is subject to protocol approval as per CPRD's guidelines. Authors are unable to share the data in its raw form, however all analytical code and codelists are open-source and found via the following links: https://github.com/flozoemartin/Birth-outcomes and https://github.com/flozoemartin/codelists.

https://github.com/flozoemartin/codelists

## Footnotes

1 Maternal models adjusted for year of birth, maternal age, practice-level IMD quintile (UK), maternal ethnicity (UK), maternal educational attainment (Norway and Sweden), household disposable income at the start of pregnancy (Sweden), smoking during pregnancy (UK and Sweden), previous stillbirth, maternal BMI at the start of pregnancy (Sweden), country of birth (Norway and Sweden), parity, antipsychotic and anti-seizure medication use in the 12 months before pregnancy, number of primary care consultations in the 12 months before pregnancy (UK and Norway), depression ever before the start of pregnancy, anxiety ever before the start of pregnancy

2 Sibling models adjusted for birth year, depression, anxiety, primary healthcare utilisation in the 12 months before pregnancy (UK and Norway), antipsychotic and anti-seizure medication in the 12 months prior to pregnancy (family average minus each siblings value to account for non-shared confounding between siblings), maternal age, maternal educational attainment (Norway and Sweden), household disposable income (Sweden), parity, smoking during pregnancy (UK and Sweden)

3 Paternal models adjusted for year of birth, maternal age, maternal educational attainment, household disposable income at the start of pregnancy (Sweden), smoking during pregnancy (Sweden), previous stillbirth, maternal BMI at the start of pregnancy (Sweden), country of birth, parity, antipsychotic and anti-seizure medication use in the 12 months before pregnancy, number of primary care consultations in the 12 months before pregnancy (Norway), maternal and paternal depression ever before the start of pregnancy, maternal and paternal anxiety ever before the start of pregnancy

4 Drug-specific analyses adjusted for year of birth, maternal age, practice-level IMD quintile (UK), maternal ethnicity (UK), maternal educational attainment (Norway and Sweden), household disposable income at the start of pregnancy (Sweden), smoking during pregnancy (UK and Sweden), previous stillbirth, maternal BMI at the start of pregnancy (Sweden), country of birth (Norway and Sweden), parity, antipsychotic and anti-seizure medication use in the 12 months before pregnancy, number of primary care consultations in the 12 months before pregnancy (UK and Norway), depression ever before the start of pregnancy, anxiety ever before the start of pregnancy

5 Drug-specific marginal risk adjusted for year of birth, maternal age, practice-level IMD quintile (UK), maternal ethnicity (UK), maternal educational attainment (Norway and Sweden), household disposable income at the start of pregnancy (Sweden), smoking during pregnancy (UK and Sweden), maternal BMI at the start of pregnancy (Sweden), country of birth (Norway and Sweden), parity

