## Supplementary material for "Antidepressant use during pregnancy and birth outcomes: analysis of electronic health data from the UK, Norway, and Sweden": Section S, Table S, Figure S

### Contents

#### Table contents

#### Figure contents

[Figure S12 Meta-analysis of adjusted estimates for each country for stratified preterm delivery analyses: moderate-to-late preterm (32-37 weeks’ gestation), very preterm (28-32 weeks’ gestation), and extremely preterm delivery (<28 weeks’ delivery).	28](#_Toc180682210)

### 1 Methods

## 1.1 UK

##### 1.1.1 CPRD GOLD

###### 1.1.1.1 The Pregnancy Register

The Pregnancy Register as provided by CPRD contains information on each pregnancy episode for each patient who has experienced a pregnancy in primary care (1). The dataset contains information such as estimated pregnancy start and end dates, the estimated length of the pregnancy, the outcome of the pregnancy, and if there was evidence of preterm delivery, for example. A substantial proportion of the pregnancy episodes in the Pregnancy Register have an unknown outcome; these are explained in detail elsewhere, but in short, pregnancies without a known outcome were pregnancy episodes identified by the Pregnancy Register algorithm that did not have sufficient codes affiliated with them in order to be able to ascertain what the outcome might have been. Given that these made up a significant proportion of the pregnancies in the Register, they were cleaned before exclusion. The cleaning process of the pregnancies without a known outcome involved using the secondary care linked data to find pregnancy episodes and align them within patients to unknown outcome pregnancy records in the Pregnancy Register. Using outcome data in HES, pertaining to either a loss in hospital or a delivery, we were able to salvage some outcome unknown pregnancies from exclusion by assigning them a new outcome from the secondary data. The algorithms were developed by PMD and HF, amended by FZM, using the codelists developed and approach laid out by Campbell *et al.* (2).

###### 1.1.1.2 Therapy data

The prescription data in CPRD is held within the Therapy file and consists of all prescriptions made to each patient in CPRD GOLD. In order to ascertain whether a prescription overlapped with pregnancy, we needed prescription start and end dates. Only prescription start date is available in the CPRD prescription data, thus we needed to derive the prescription end date using daily dose (the number of doses prescribed per day) and quantity (the number of doses given per prescription). These variables are provided by CPRD but require some post-hoc cleaning for unusual and missing values. Daily dose was supplemented with dose number multiplied by dose frequency and quantity was supplemented with pack size, where available Where some or all of these values were missing, we used hot-decking imputation to fill in the gaps in the prescription data. The approach follows a modal approach with decreasing levels of specificity until all the gaps are filled: first, missing daily dose and quantity are filled in using the modal value of each variable from prescriptions of the same drug within the same patient. If no such data are available, then the modal value from the same prescriptions among all patients are taken, and so on until the least specific criteria, among the same class, is used to fill in the final missing values.

All antidepressants that fall under the N06A WHO ATC classification were investigated, except bupropion given that it’s use is not indicated for depression in the UK (Supplement S1).

Dose was derived for each drug, by taking the dose prescribed per day and presented as daily dose in milligrams. Given that different medications are dosed differently, but all follow thresholds of low, medium, and high dosing, we wanted to standardize doses across each medication for comparability in the secondary analysis of dose.

To standardize dose, we plotted to distribution of dose for each medication and identified the dose value that represented each quartile of the distribution. We assigned low dose as the bottom 25% of the distribution, medium as the middle 50% of the distribution, and high as the top 75%. For drugs that are rarely prescribed or have few dose values that are prescribed in clinical practice, these thresholds had to be amended to more stringent upper and lower bounds, for example agomelatine required an upper threshold of 90% to capture true high dose in the high dose category. The spreadsheet showing the dose thresholds for each medication is attached in the Appendix. The doses assigned to each medication were checked for clinical agreement in EMC and with clinical coauthors.

##### 1.1.2 Linked data

###### 1.1.2.1 Hospital Episode Statistic Maternity

CPRD GOLD has several secondary care linked data sources for the majority of English practices (around 75%) (3). Within the Hospital Episode Statistics (HES) Admitted Patient Care data, HES Maternity data contains a range of different delivery and birth-related measures, including birthweight, gestational length, child sex, and more. The data in HES Maternity is divided into delivery records (affiliated with the patient who is giving birth) and birth records (the patient who being born), so can be linked to maternal or baby patient identifiers depending on whose record it is.

Records pertaining to the same delivery were combined by episode date and all available information on birthweight and child sex were compared and retained if consistent across records.

#### 1.2 Norway

In the Nordic countries, including Norway, detailed medical information on every resident (with a personal identifier (PIN) number) is collected into a series of compulsory, population-based registries (4). The registries consist of the Norwegian Patient Registry (NPR), the Medical Birth Registry of Norway (MBRN), and the Norwegian Prescription Database (NorPD), among others. These registries have been harmonized across the Nordic countries for pregnancy pharmacoepidemiology research into the Nordic Pregnancy Drug Safety Studies (NorPreSS) common data model (CDM).

##### 1.2.1 The Norwegian Patient Registry (NPR)

NPR was established in 2008 and collects data from specialist health services, making it one of the largest registries in Norway (5). It contains medical information such as diagnostic codes, procedure codes, and dates pertaining to each medical event (6). Diagnostic codes are either International Statistical Classification of Diseases and Related Health Problems (ICD)-10 codes (7). Depression and anxiety were obtained from these data.

##### 1.2.2  The Norwegian Control and Payment of Health Reimbursement Database (KUHR)

KUHR was established in 2006 and contains diagnoses and dates pertaining to each medical event made in primary care in Norway (8). Diagnoses in KUHR are coded using International Classification of Primary Care (ICPC)-2 codes. Depression, anxiety, and number of primary care consultations were obtained from these data.

##### 1.2.3 The Medical Birth Registry of Norway

The MBRN was set up in 1967 following the thalidomide tragedy to initiate surveillance of birth defects and ultimately prevent future harm by providing a platform for research (9). Data on all pregnancies after 12 weeks’ gestation in Norway are provided by attending midwives and obstetricians, including maternal health before and during pregnancy, pregnancy and delivery complications, and information about the infant (9).

##### 1.2.4 The Norwegian Prescription Database

NorPD was established in 2004 with aims to describe patterns of drug prescribing in Norway, promote research into drug safety and efficacy, manage quality of prescribing, and act as internal controls for prescribing physicians (10). It contains pseudonymised patient and prescriber identifiers and information on the drug prescribed like Defined Daily Dose (DDD) and Anatomical Therapeutic Chemical (ATC) codes, as described in detail by the World Health Organisation Collaborating Centre (WHOCC) for Drug Statistics Methodology (11).

To identify patients who were dispensed antidepressants during pregnancy, the total DDD (analogous to duration of a treatment) was added to the dispensation date to estimate the treatment end date. A pregnancy was coded as “exposed to antidepressants” if there were any antidepressant dispensations made during pregnancy or prescriptions that overlapped with pregnancy.

##### 1.2.5 Statistics Norway

For high quality epidemiological studies of health-related exposures and outcomes, consideration of factors outside of the data collected within the medical registries is imperative. Statistics Norway is responsible for the collection of data pertaining to economic factors, including educational attainment and migration, from a local to a national level (12). Educational attainment was obtained from these data.

#### 1.3 Sweden’s registries

Similarly to Norway, information is collected on every resident (with a unique personal number) in Sweden from their medical, tax, and education-related records making up extensive, population-based registries. The registries include but are not limited to the Swedish National Patient Register (NPR), the Medical Birth Registry of Sweden (MBR), and the Swedish Prescribed Drug Register (PDR). These data were obtained via the DOHaD project at Karolinska Institutet and harmonised with the Norwegian and UK data.

##### 1.3.1 The National Patient Registry

The Swedish NPR was established in the 1960’s and contains information on inpatient stays from 1964, with full population coverage by 1987 (13). It contains information on hospital stays, treatment administered in specialist care, and more recently information on compulsory psychiatric care (14). Diagnostic codes are International Statistical Classification of Diseases and Related Health Problems (ICD)-9 or -10 codes, depending on the age of the record (ICD-9 pre-1997) (14).

##### 1.3.2 The Medical Birth Registry of Sweden

The MBR was set up in 1973 to capture every birth in Sweden and today, has about 98% coverage of the Swedish births. It’s pseudonymisation process is similar to Norway and the use of personal numbers allow for wide linkage across other Swedish registries (15). The MBR is intended to cover all live births, regardless of gestational week, and all stillbirths ≥22 weeks’ gestation (≥28 weeks’ gestation before June 2008) (16). Included in the MBR are the self-reported medications semi-automatically captured from free text from the first antenatal appointment (16), and between 1995 and July 2005 was the only source of information regarding medication use during pregnancy in Sweden.

##### 1.3.3 The Swedish Prescribed Drug Register

The Swedish PDR was established in July 2005 and contains information on all dispensed drugs in ambulatory settings (i.e., excluding those prescribed during hospital admission) to individuals on a national level (17). It is organised by ATC code and total DDD, similarly to Norway and described above (11). The Swedish PDR supplements the self-reported medication use during pregnancy as recorded antenatally, organised by ATC code, from mid-2005.

Similarly to the Norwegian PDR, the total DDD (analogous to duration of prescription) was added to the dispensation date to estimate the prescription end date. A pregnancy was coded as “exposed to antidepressants” if there were any antidepressant dispensations made during pregnancy or prescriptions that overlapped with pregnancy.

##### 1.3.4 Statistics Sweden

Statistics Sweden collects data on income and tax history (18), into registers such as the Longitudinal Integrated Database for Health Insurance and Labour Market Studies (LISA). These are important variables allowing high quality epidemiological studies to be performed adjusting for various confounders.

##### 1.3.5 Developmental Origins of Health And Disease

The Swedish registry data was compiled in part for me by Michael Lundberg at Karolinska Institutet. I received a dataset containing a mix of data from the Swedish NPR pertaining to maternal and paternal indications and data regarding pregnancy and birth information from the Swedish MBR.

#### 1.4 Antidepressant codes

Codes pertaining to each included medication: ATC codes applied to the Norwegian and Swedish dispensation data and BNF codes (matched to CPRD GOLD ‘medcodes’) in CPRD GOLD’s prescription data and whether these drugs were included in the ‘other’ category in each country.

Table S1 Antidepressant codes used to define the exposure in the present study.

| **GROUP** | **ATC code** | **Name** | **CPRD prodcode** |
| --- | --- | --- | --- |
| N06AA | N06AA01 | desipramine | 7981, 7979 |
| N06AA | N06AA02 | imipramine | 3668, 7573, 7784, 10649, 1310, 41681, 34222, 67935, 71253, 70287, 32863, 34872, 81638, 1809, 34813, 34355, 82911, 83472, 80885, 41408, 8055, 42247, 33074, 2579, 56501, 7910, 4404 |
| N06AA | N06AA03 | imipramine oxide |  |
| N06AA | N06AA04 | clomipramine | 7515, 3657, 8719, 7693, 7894, 3194, 34866, 68665, 83468, 41628, 62620, 43561, 3670, 34245, 81617, 83470, 41563, 45350, 65762, 8720, 64458, 3925, 45318, 41597, 53187, 78324, 65804, 77227, 53161, 38274, 78057, 8661, 3195, 79276, 25036 |
| N06AA | N06AA05 | opipramol |  |
| N06AA | N06AA06 | trimipramine | 8928, 2532, 2531, 4310, 42228, 53808, 2039, 45226, 57978, 66493,3196, 65445, 66919, 65213 |
| N06AA | N06AA07 | lofepramine | 25070, 58450, 2093, 41627, 114, 34046, 34950, 71067, 74586, 66100, 34578, 68657, 67742, 56703, 34672, 60591, 56229, 43534, 4218, 77717, 25444, 79397 |
| N06AA | N06AA08 | dibenzepin |  |
| N06AA | N06AA09 | amitriptyline | 34916, 45242, 83, 33090, 52867, 24141, 57972, 55491, 70991, 61835, 76839, 45233, 34731, 66578, 83079, 80135, 57107, 65879, 24152, 59161, 34401, 46801, 22070, 64000, 79826, 70300, 77229, 76298, 46818, 3777, 76927, 487, 34197, 41729, 42394, 34474, 32439, 49, 34782, 54877, 24145, 55139, 42078, 71042, 65987, 64647, 79766, 34503, 24134, 66579, 60355, 77167, 65439, 66572, 24147, 34129, 6312, 83377, 78364, 67127, 34224, 60410, 83516, 4682, 40396, 1888, 34274, 34634, 64330, 82403, 78221, 46970, 34182, 69712, 33624, 34107, 4690, 34251, 59820, 64141, 76952, 2525, 77497, 48065, 26213, 20026, 27008, 24680, 2486, 2985, 8726, 8878, 7751, 8332, 8831, 27876, 873, 79401, 13496, 23497, 3771, 30738, 8250, 20712 |
| N06AA | N06AA10 | nortriptyline | 12194, 7677, 8640, 83498, 17183, 12549, 12353, 4118, 3183, 65237, 80352, 55970, 39145, 72626, 80546, 68228, 7678, 3903, 48216, 63276, 66201, 78224, 83536, 69317 |
| N06AA | N06AA11 | protriptyline | 4149, 7755, 7816, 11187, 7756, 60929 |
| N06AA | N06AA12 | Doxepin | 3842, 3554, 40777, 5073, 73363, 81337, 7059, 35258, 35493, 10413, 12129, 12125, 14519 |
| N06AA | N06AA13 | Iprindole | 27476, 27733, 24700, 31672 |
| N06AA | N06AA14 | Melitracen |  |
| N06AA | N06AA15 | Butriptyline | 12227, 32457, 18932 |
| N06AA | N06AA16 | Dosulepin | 43024, 77130, 70838, 84, 23426, 34745, 34643, 31824, 44853, 29875, 33164, 34641, 76317, 34223, 19168, 50722, 45737, 6054, 71023, 70593, 74, 32121, 19186, 67728, 42734, 81126, 31826, 34525, 62681, 71059, 34058, 57926, 10948, 1940, 15632, 21820, 21819, 67990, 51758, 1169, 2320, 30376, 21157 |
| N06AA | N06AA17 | amoxapine | 3652, 4411, 17319, 3351, 21357, 24723, 15380, 14398, 55289, 7475 |
| N06AA | N06AA18 | Dimetacrine |  |
| N06AA | N06AA19 | Amineptine |  |
| N06AA | N06AA21 | Maprotiline | 20379, 12222, 10645, 14410, 13558, 16727, 8549, 12100, 20405 |
| N06AA | N06AA23 | Quinupramine |  |
| N06AB | N06AB02 | Zimeldine |  |
| N06AB | N06AB03 | Fluoxetine | 79506, 79494, 33071, 67431, 69941, 77881, 42499, 81906, 75645, 38890, 22, 19183, 71852, 45329, 60962, 75799, 67736, 45247, 75688, 34202, 34294, 69525, 59358, 66744, 34288, 42107, 62155, 19470, 45224, 67769, 34456, 34849, 67092, 45316, 33410, 60534, 60138, 2548, 34216, 42803, 60619, 73414, 30258, 36893, 68266, 69685, 74886, 67496, 79590, 67562, 75068, 78889, 4075, 75247, 67888, 34856, 62335, 14740, 67758, 77381, 418, 48220, 61335, 69542, 57532, 252, 80702, 75943, 4907, 37256, 33779, 29786 |
| N06AB | N06AB04 | Citalopram | 815, 513, 57936, 56292, 72124, 3861, 79784, 63953, 1712, 2408, 34498, 476, 34586, 64423, 32848, 49165, 42660, 52100, 59650, 53787, 71005, 33720, 52408, 34436, 45286, 75697, 52824, 59193, 63441, 34499, 60888, 41528, 80370, 56355, 34413, 54827, 34722, 67, 34356, 67097, 34871, 53394, 48026, 56009, 58476, 52607, 52354, 34415, 34970, 81067, 73417, 72373,26016, 82670, 34966, 60568, 34822, 71848, 74753, 43519, 4770, 36746, 69571, 46977, 75075, 60839, 70790, 55033, 75702, 83078, 34603, 45223, 34466, 45304, 46926, 32546, 29756 |
| N06AB | N06AB05 | Paroxetine | 35021, 76946, 59288, 81854, 67259, 527, 50, 34419, 32899, 73668, 40892, 34351, 55023, 33978, 79383, 79381, 1397, 34587, 40165, 64785, 83463, 78843, 68325, 75054, 35112, 66292, 74588, 841, 73589, 77650, 82506, 3601, 1575, 55537, 76772, 82578 |
| N06AB | N06AB06 | Sertraline | 82495, 82384, 4352, 77385, 82497, 1612, 727, 55146, 62950, 61503, 59600, 62692, 69726, 67928, 66560, 54933, 66413, 68756, 44944, 73962, 49519, 77607, 78278, 62819, 54826, 78626, 73759, 82884, 83101, 82195, 54081, 83668, 488, 32401, 58723, 42387, 45915, 62693, 69725, 63481, 58664, 67730, 81991, 69898, 55488, 75952, 62927, 75405, 7328, 77538, 77707, 65771 |
| N06AB | N06AB07 | Alaproclate |  |
| N06AB | N06AB08 | Fluvoxamine | 12123, 2897, 2290, 48045, 44861, 43518, 2880 |
| N06AB | N06AB09 | Etoperidone |  |
| N06AB | N06AB10 | Escitalopram | 74785, 648, 83378, 74858, 26056, 80820, 6360, 80821, 41062, 785, 603, 81987, 63916, 74993, 82346, 20152, 6218, 80453, 72773, 40726, 6405 |
| N06AF | N06AF01 | Isocarboxazid | 41731, 12207, 12503 |
| N06AF | N06AF02 | Nialamide |  |
| N06AF | N06AF03 | Phenelzine | 3349, 82839, 4321 |
| N06AF | N06AF04 | Tranylcypromine | 10787, 3783, 41654 |
| N06AF | N06AF05 | Iproniazide | 25945, 18290 |
| N06AF | N06AF06 | Iproclozide |  |
| N06AG | N06AG02 | Moclobemide | 9206, 5832, 2883, 67305, 41747, 5187 |
| N06AG | N06AG03 | Toloxatone |  |
| N06AX | N06AX01 | Oxitriptan |  |
| N06AX | N06AX02 | Tryptophan | 3353, 54747, 5611, 20504, 12221, 54686, 4422 |
| N06AX | N06AX03 | Mianserin | 7468, 8144, 8585, 3083, 47363, 4329, 6255, 12368, 11956, 12192 |
| N06AX | N06AX04 | Nomifensine |  |
| N06AX | N06AX05 | Trazodone | 81245, 81210, 4194, 4003, 4874, 8174, 13621, 1730, 34580, 73639, 19181, 41709, 41710, 65152, 72291, 66749, 12710, 4020, 73419, 77915, 73636, 76480, 30983, 29857, 34470, 55137, 55138, 83013, 57226, 3355, 34003, 71031, 29339, 41609, 34421, 61842, 6442, 59931, 70521, 77474, 80399, 61657, 69355 |
| N06AX | N06AX06 | Nefazodone | 3391, 4297, 63827, 4554, 4011, 67757, 9534 |
| N06AX | N06AX07 | Minaprine |  |
| N06AX | N06AX08 | Bifemelane |  |
| N06AX | N06AX09 | Viloxazine |  |
| N06AX | N06AX10 | Oxaflozane |  |
| N06AX | N06AX11 | Mirtazapine | 6421, 43253, 64101, 43241, 66580, 61856, 43248, 43246, 68680, 55482, 58291, 77865, 65555, 43237, 48698, 54012, 6795, 43239, 53699, 66183, 59953, 46668, 66752, 82469, 43242, 54342, 82206, 54644, 82682, 74557, 43257, 16154, 53321, 81914, 61547, 47966, 68544, 6488, 43250, 53648, 68052, 48185, 69420, 76187, 59694, 80953, 742, 47945, 40160, 54792, 69005, 77488, 78654, 60538, 82070, 82326, 56209, 68933, 71543, 63403, 6481, 43235, 43236, 43256, 43247, 64139, 43234, 49820, 6854, 33337, 58625, 59954, 64223, 82245, 77377, 4726, 67272, 60370, 6846, 50892, 82520, 80893, 10083, 53543, 82510, 15268, 31168, 79008, 79252 |
| N06AX | N06AX13 | Medifoxamine |  |
| N06AX | N06AX14 | Tianeptine |  |
| N06AX | N06AX15 | Pivagabine |  |
| N06AX | N06AX16 | Venlafaxine | 52516, 52074, 71806, 61236, 45664, 45959, 65738, 67271, 623, 6274, 67288, 77089, 9182, 81253, 74010, 5710, 51280, 65899, 74011, 75894, 1474, 76771, 81158, 43968, 80721, 83355, 43673, 41299, 48199, 41314, 41033, 59753, 60843, 40817, 40815, 39809, 39770, 57751, 52716, 40514, 40515, 70420, 70495, 69819, 70315, 50081, 59035, 49511, 58726, 74516, 58681, 55424, 55501, 2654, 70806, 60549, 71782, 43334, 39360, 50934, 62734, 65666, 40054, 80331, 58837, 45806, 301, 56662, 73667, 68050, 75525, 59923, 70353, 83069, 51361, 60895, 51699, 13237, 2617, 470, 71257, 59563, 68876, 43203, 39359, 1222, 60449, 73658, 66437, 56457, 63859, 53326, 63268, 40062, 40407, 80541, 45818, 40059, 44936, 44937, 71932, 70931, 40092, 67563, 40277, 7672, 75263, 40517, 42600, 40764, 40917, 40049, 78585, 40048, 75848 |
| N06AX | N06AX17 | Milnacipran |  |
| N06AX | N06AX18 | Reboxetine | 15163, 2356 |
| N06AX | N06AX19 | Gepirone |  |
| N06AX | N06AX21 | Duloxetine | 13151, 82572, 14849, 63216, 82860, 73298, 73540, 82903, 74190, 7153, 70063, 67564, 65165, 82340, 71669, 7122, 65618, 65809, 70728, 80840, 74907, 66412, 79628, 63370, 62688, 70405, 82035, 81423, 76857, 7147, 79768, 82430, 6895, 63763, 65888, 69428, 73868, 69965, 69752, 66405, 72211, 65892, 51383, 64442, 78777, 68096, 74774, 16969, 14803, 79275 |
| N06AX | N06AX22 | Agomelatine | 40494  40295 |
| N06AX | N06AX23 | Desvenlafaxine |  |
| N06AX | N06AX24 | Vilazodone |  |
| N06AX | N06AX25 | Hyperici herba |  |
| N06AX | N06AX26 | Vortioxetine | 67874, 69991, 69992, 65483, 66890, 65482 |
| N06AX | N06AX27 | Esketamine |  |
| N06AX | N06AX28 | Levomilnacipran |  |
| N06AX | N06AX29 | Brexanolone |  |

#### 1.5 Covariate selection

Table S2 Covariates that were included in the primary adjustment set based on *a priori* knowledge and ≤5% missing data.

| **Covariate** | **Included in primary adjustment set (≤5% missing data?)** | | |
| --- | --- | --- | --- |
|  | **UK** | **Norway** | **Sweden** |
| Year of birth for the pregnancy | ✔ | ✔ | ✔ |
| Maternal age at delivery | ✔ | ✔ | ✔ |
| Maternal educational attainment at start of pregnancy | - | ✔ | ✔ |
| Maternal practice-level IMD quintile | ✔ | - | - |
| Maternal body mass index around the start of pregnancy | **✗** | **✗** | ✔ |
| Maternal country of birth | - | ✔ | ✔ |
| Maternal ethnicity | ✔ | - | - |
| Maternal parity | ✔ | ✔ | ✔ |
| Previous stillbirth | ✔ | ✔ | ✔ |
| Maternal anti-seizure medication (ASM) use in the 12 months before pregnancy | ✔ | ✔ | ✔ |
| Maternal antipsychotic (AP) medication use in the 12 months before pregnancy | ✔ | ✔ | ✔ |
| Maternal smoking around the start of pregnancy | ✔ | **✗** | ✔ |
| Maternal depression before the start of pregnancy | ✔ | ✔ | ✔ |
| Maternal anxiety before the start of pregnancy | ✔ | ✔ | ✔ |

### Results

#### Study population

Figure S1 Flow diagrams of pregnancies through the study in each country: UK, Norway, and Sweden.

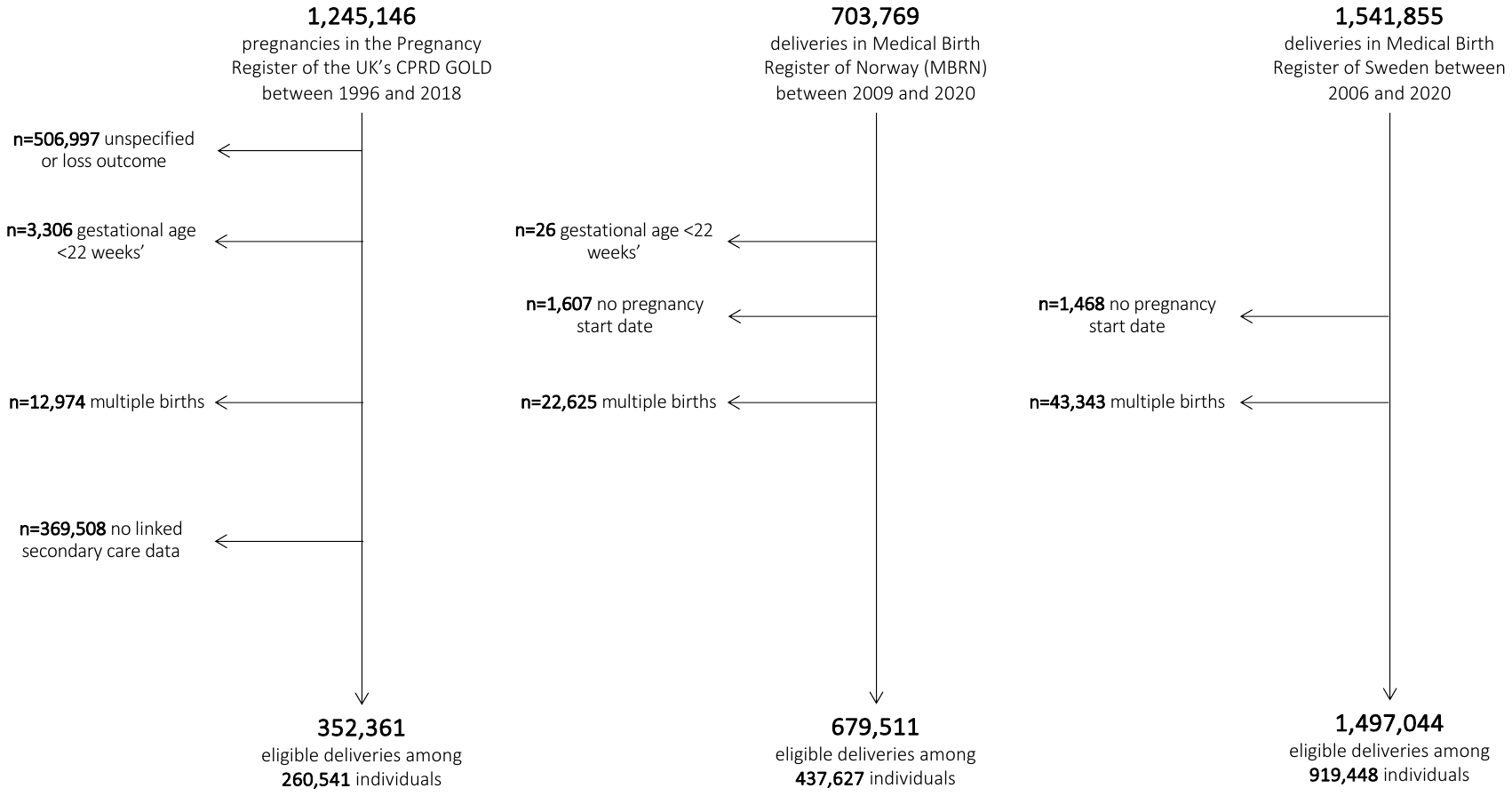

Table S3 Proxy measures of socioeconomic position in each country.

|  | **UK** | | | **Norway** | | **Sweden** | |
| --- | --- | --- | --- | --- | --- | --- | --- |
|  | **Exposed to ADs** | | **Unexposed to ADs** | **Exposed to ADs** | **Unexposed to ADs** | **Exposed to ADs** | **Unexposed to ADs** |
| **Maternal educational attainment at the start of pregnancy** | | | | | | | |
| Compulsory or less | - | - | | 5,228 (30.4) | 112,508 (17.0) | 35,075 (43.1) | 572,936 (40.5) |
| Secondary | - | - | | 5,166 (30.0) | 160,747 (24.3) | 27,025 (33.2) | 548,849 (38.8) |
| Post-secondary | - | - | | 5,021 (29.2) | 247,013 (37.3) | 10,683 (13.1) | 150,570 (10.6) |
| Post-graduate | - | - | | 1,514 (8.8) | 111,484 (16.8) | 7,973 (9.8) | 109,291 (7.7) |
| Missing (likely educated abroad) | - | - | | 273 (1.6) | 30,557 (4.6) | 586 (0.7) | 34,056 (2.4) |
| **Household disposable income at the start of pregnancy (in quintiles)** | | | | | | | |
| 1 | - | - | | - | - | 11,766 (14.5) | 249,062 (17.6) |
| 2 | - | - | | - | - | 19,121 (23.5) | 271,848 (19.2) |
| 3 | - | - | | - | - | 18,433 (22.7) | 287,247 (20.3) |
| 4 | - | - | | - | - | 16,678 (20.5) | 297,205 (21.0) |
| 5 | - | - | | - | - | 15,329 (18.8) | 309,711 (21.9) |
| **Practice-level Index of Multiple Deprivation score (in quintiles)** | | | | | | | |
| 1 (least deprived) | 2,550 (11.8) | 52,135 (15.8) | | - | - | - | - |
| 2 | 3,460 (16.0) | 57,493 (17.4) | | - | - | - | - |
| 3 | 4,013 (18.5) | 60,791 (18.4) | | - | - | - | - |
| 4 | 4,904 (22.6) | 71,051 (21.5) | | - | - | - | - |
| 5 (most deprived) | 6,738 (31.1) | 89,226 (27.0) | | - | - | - | - |
| **Maternal country of birth** | | | | | | | |
| Foreign-born | - | - | | 2,678 (15.6) | 180,878 (27.3) | 9,687 (11.9) | 384,636 (27.2) |
| Missing | - | - | | 122 (0.7) | 3,900 (0.6) | 55 (0.1) | 1,113 (0.1) |
| **Maternal ethnicity** | | | | | | | |
| White | 20,389 (94.1) | 289,532 (87.6) | | - | - | - | - |
| South Asian | 469 (2.2) | 16,902 (5.1) | | - | - | - | - |
| Black | 245 (1.1) | 8,221 (2.5) | | - | - | - | - |
| Other | 143 (0.7) | 5,449 (1.6) | | - | - | - | - |
| Mixed | 200 (0.9) | 3,180 (1.0) | | - | - | - | - |
| Missing | 219 (1.0) | 7,412 (2.2) | | - | - | - | - |

**Table S4** Outcome prevalence stratified by antidepressant exposure during pregnancy in the UK, Norway, and Sweden.

| **Outcome** | **UK** | | **Norway** | | **Sweden** | | |
| --- | --- | --- | --- | --- | --- | --- | --- |
|  | **Exposed to ADs** | **Unexposed to ADs** | **Exposed to ADs** | **Unexposed to ADs** | **Exposed to ADs** | | **Unexposed to ADs** |
| **Total** | 21,665 (100.0) | 330,696 (100.0) | 17,202 (100.0) | 662,309 (100.0) | 81,342 (100.0) | 1,415,702 (100.0) | |
| **Birth outcome** |  |  |  |  |  |  | |
| Stillborn | 121 (0.6) | 1,506 (0.5) | 69 (0.4) | 2,139 (0.3) | 355 (0.4) | 4,842 (0.3) | |
| Missing | 0 (0) | 0 (0) | 0 (0) | 0 (0) | 0 (0) | 0 (0) | |
| Neonatal death | - | - | 34 (0.2) | 878 (0.1) | 123 (0.2) | 1,846 (0.1) | |
| Missing | - | - | 69 (0.4) | 2,139 (0.3) | 0 (0) | 0 (0) | |
| Apgar score < 7 at 5 mins | - | - | 478 (2.8) | 10,635 (1.6) | 2,576 (3.2) | 20,361 (1.4) | |
| Missing | - | - | 12 (0.1) | 432 (0.1) | 594 (0.7) | 8,233 (0.6) | |
| **Gestational age** |  |  |  |  |  |  | |
| Preterm delivery (<37 weeks') | 1,869 (8.6) | 21,805 (6.6) | 1,270 (7.4) | 32,190 (4.9) | 5,901 (7.3) | 65,955 (4.7) | |
| Term delivery (37-42 weeks') | 18,783 (86.7) | 292,182 (88.4) | 15,402 (89.5) | 599,993 (90.6) | 72,144 (88.7) | 1,254,933 (88.6) | |
| Post-term delivery (>42 weeks') | 1,013 (4.7) | 16,709 (5.1) | 530 (3.1) | 30,126 (4.5) | 3,297 (4.1) | 94,814 (6.7) | |
| **Total with birthweight and sex** | 16,343 (75.4) | 239,799 (72.5) | 17,198 (100.0) | 662,234 (100.0) | 81,342 (100.0) | 1,415,702 (100.0) | |
| **Size for gestational age (percentile distribution)** |  |  |  |  |  |  | |
| Small for gestational age (SGA, <10th percentile) | 2,049 (12.5) | 23,198 (9.7) | 1,766 (10.3) | 65,372 (9.9) | 7,707 (9.5) | 140,496 (9.9) | |
| Adequate for gestational age (AGA, 10-90th percentile) | 12,806 (78.4) | 192,874 (80.4) | 13,754 (80.0) | 533,395 (80.5) | 64,551 (79.4) | 1,134,039 (80.1) | |
| Large for gestational age (LGA, >90th percentile) | 1,488 (9.1) | 23,727 (9.9) | 1,678 (9.8) | 63,467 (9.6) | 8,968 (11.0) | 139,564 (9.9) | |
| **Size for gestational age (INTERGROWTH-21)** |  |  |  |  |  |  | |
| SGA | 1,636 (10.0) | 17,917 (7.5) | 676 (3.9) | 23,388 (3.5) | 2,944 (3.6) | 51,284 (3.6) | |
| AGA | 12,545 (76.8) | 187,331 (78.1) | 12,437 (72.3) | 481,669 (72.7) | 57,662 (70.9) | 1,018,022 (71.9) | |
| LGA | 3,446 (21.1) | 56,818 (23.7) | 4,089 (23.8) | 157,252 (23.7) | 20,736 (25.5) | 346,396 (24.5) | |
| **Size for gestational age (new Swedish standard)** |  |  |  |  |  |  | |
| SGA | 4,699 (28.8) | 60,648 (25.3) | 3,656 (21.3) | 133,668 (20.2) | 12,657 (15.6) | 228,340 (16.1) | |
| AGA | 11,493 (70.3) | 178,033 (74.2) | 12,724 (74.0) | 499,025 (75.4) | 61,045 (75.0) | 1,083,804 (76.6) | |
| LGA | 1,435 (8.8) | 23,390 (9.8) | 822 (4.8) | 29,616 (4.5) | 7,640 (9.4) | 103,558 (7.3) | |

Table S5 Proportion of preterm that fall into moderate-to-late, very, and extremely preterm categories.

| **Types of preterm delivery** | **UK** | | **Norway** | | **Sweden** | |
| --- | --- | --- | --- | --- | --- | --- |
|  | **Exposed to ADs** | **Unexposed to ADs** | **Exposed to ADs** | **Exposed to ADs** | **Unexposed to ADs** | **Exposed to ADs** |
| Total preterm | 1,869 (100) | 21,805 (100) | 1,270 (100) | 32,190 (100) | 5,901 (100) | 65,955 (100) |
| Moderate-to-late preterm (32-37 weeks') | 1,533 (82.0) | 17,479 (80.2) | 1,091 (85.9) | 26,907 (83.6) | 5,135 (87.0) | 54,953 (83.3) |
| Very preterm (28-32 weeks') | 228 (12.2) | 2,907 (13.3) | 107 (8.4) | 3,119 (9.7) | 470 (8.0) | 6,489 (9.8) |
| Extremely preterm (<28 weeks') | 108 (5.8) | 1,419 (6.5) | 72 (5.7) | 2,164 (6.7) | 296 (5.0) | 4,513 (6.8) |

#### 1.6 Maternal analysis

Figure S2 Individual country and pooled odds ratios from fixed-effect meta-analysis of maternal models^[1]^ (any antidepressant use during pregnancy compared with no use) for each outcome in each outcome where available.

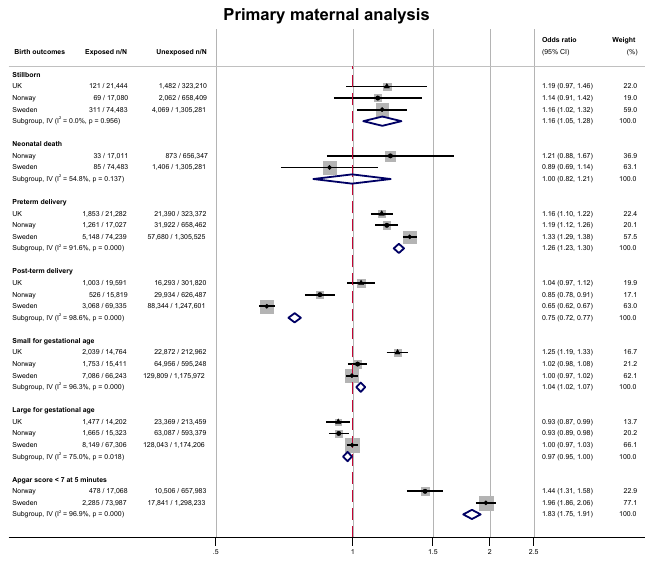

##### Trimester-specific analysis

Figure S3 Individual and pooled odds ratios from fixed effect meta-analyses of antidepressant use in each trimester^[2]^ in the UK, Norway and Sweden for each outcome.

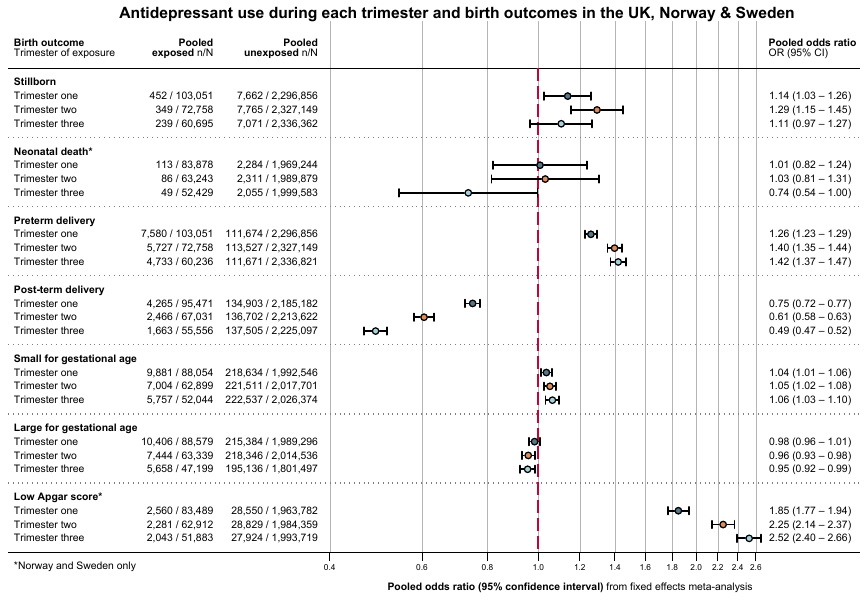

##### Drug-specific analyses

Figure S4 Individual and pooled odds ratios from fixed effect meta-analyses of individual antidepressants during pregnancy and preterm delivery^[3]^ in the UK, Norway and Sweden for each outcome.

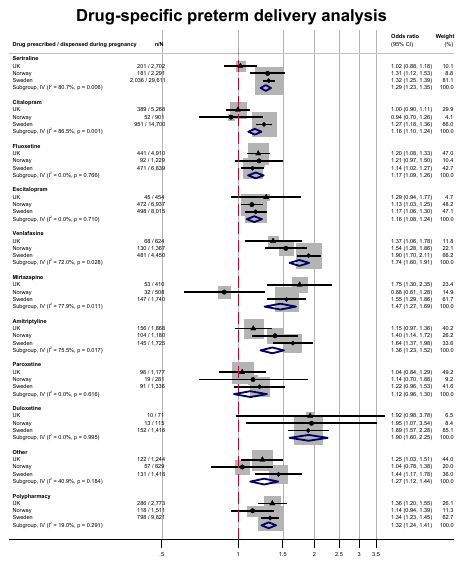

Table S6 Drug-specific analysis of gestational age.

|  | **Gestational age** | | | | | | | | | | | |
| --- | --- | --- | --- | --- | --- | --- | --- | --- | --- | --- | --- | --- |
|  | **UK** | | | | **Norway** | | | | **Sweden** | | | |
|  | **Total** | **Marginal mean gestational age** | **Adjusted^[4]^ mean difference (95%CI)** | **P-value** | **Total** | **Marginal mean gestational age** | **Adjusted mean difference (95%CI)** | **P-value** | **Total** | **Marginal mean gestational age** | **Adjusted mean difference (95%CI)** | **P-value** |
| Unexposed | 330,860 | 39.33 | 0 (ref) | - | 662,362 | 39.37 | 0 (ref) | - | 1,305,525 | 39.42 | 0 (ref) | - |
| Sertraline exposed | 2,702 | 39.23 | -0.10 (-0.18, -0.02) | 0.014 | 2,291 | 39.15 | -0.22 (-0.31, -0.13) | 0.000 | 27,220 | 39.09 | -0.33 (-0.35, -0.30) | 0.000 |
| Citalopram exposed | 5,268 | 39.32 | -0.01 (-0.07, 0.05) | 0.646 | 901 | 39.28 | -0.09 (-0.22, 0.03) | 0.144 | 13,515 | 39.14 | -0.28 (-0.31, -0.24) | 0.000 |
| Fluoxetine exposed | 4,910 | 39.24 | -0.09 (-0.16, -0.03) | 0.006 | 1,229 | 39.10 | -0.27 (-0.39, -0.16) | 0.000 | 6,236 | 39.15 | -0.27 (-0.32, -0.22) | 0.000 |
| Escitalopram exposed | 454 | 39.23 | -0.11 (-0.32, 0.11) | 0.344 | 6,937 | 39.25 | -0.12 (-0.17, -0.07) | 0.000 | 7,425 | 39.21 | -0.21 (-0.25, -0.17) | 0.000 |
| Venlafaxine exposed | 624 | 39.06 | -0.27 (-0.46, -0.08) | 0.005 | 1,367 | 39.00 | -0.37 (-0.48, -0.26) | 0.000 | 4,040 | 38.83 | -0.59 (-0.66, -0.52) | 0.000 |
| Mirtazapine exposed | 410 | 39.06 | -0.27 (-0.51, -0.03) | 0.029 | 508 | 39.39 | 0.02 (-0.16, 0.19) | 0.861 | 1,571 | 39.17 | -0.25 (-0.36, -0.14) | 0.000 |
| Amitriptyline exposed | 1,868 | 39.21 | -0.12 (-0.22, -0.02) | 0.018 | 1,180 | 39.09 | -0.28 (-0.41, -0.15) | 0.000 | 1,572 | 39.03 | -0.39 (-0.49, -0.29) | 0.000 |
| Paroxetine exposed | 1,177 | 39.23 | -0.10 (-0.23, 0.04) | 0.164 | 281 | 39.29 | -0.08 (-0.32, 0.16) | 0.503 | 1,206 | 39.19 | -0.23 (-0.34, -0.12) | 0.000 |
| Duloxetine exposed | 71 | 38.64 | -0.69 (-1.34, -0.05) | 0.035 | 115 | 38.92 | -0.45 (-0.91, 0.00) | 0.051 | 1,266 | 38.84 | -0.58 (-0.70, -0.46) | 0.000 |
| Other exposed^[5]^ | 1,244 | 39.13 | -0.21 (-0.34, -0.07) | 0.003 | 829 | 39.37 | -0.01 (-0.14, 0.13) | 0.933 | 1,268 | 39.18 | -0.24 (-0.37, -0.12) | 0.000 |
| Polypharmacy exposed^[6]^ | 2,773 | 39.16 | -0.17 (-0.26, -0.09) | 0.000 | 1,511 | 39.12 | -0.25 (-0.36, -0.15) | 0.000 | 8,920 | 39.00 | -0.41 (-0.46, -0.37) | 0.000 |

Figure S5 Individual and pooled odds ratios from fixed effect meta-analyses of individual antidepressants during pregnancy and SGA^[7]^ in the UK, Norway and Sweden for each outcome.

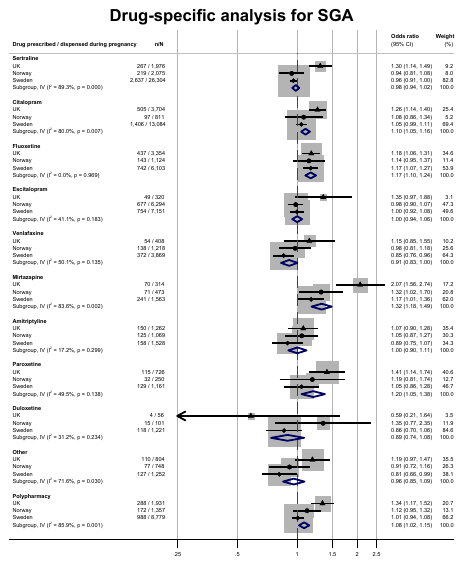

Table S7 Drug-specific analysis of birthweight and small for gestational age in UK (average birth weight 3,330g).

|  | **Birth weight (in grams)** | | | | | | | | | | | |
| --- | --- | --- | --- | --- | --- | --- | --- | --- | --- | --- | --- | --- |
|  | **UK** | | | | **Norway** | | | | **Sweden** | | | |
|  | **Total** | **Marginal mean gestational age** | **Adjusted^[8]^ mean difference (95%CI)** | **P-value** | **Total** | **Marginal mean gestational age** | **Adjusted mean difference (95%CI)** | **P-value** | **Total** | **Marginal mean gestational age** | **Adjusted mean difference (95%CI)** | **P-value** |
| Unexposed | 216,072 | 3,398 | 0 (ref) | - | 598,767 | 3525.25 | 0 (ref) | - | 1,175,972 | 3539 | 0 (ref) | - |
| ()Sertraline exposed | 1,976 | 3,349 | -48 (-73, -24) | 0.000 | 2,075 | 3492.19 | -33.06 (-59, -8) | 0.011 | 24,322 | 3493 | -46 (-54, -39) | 0.000 |
| Citalopram exposed | 3,704 | 3,377 | -21 (-39, -2) | 0.028 | 811 | 3504.83 | -20.41 (-60, 19) | 0.307 | 12,107 | 3481 | -58 (-68, -48) | 0.000 |
| Fluoxetine exposed | 3,354 | 3,352 | -45 (-65, -26) | 0.000 | 1,124 | 3449.89 | -75.36 (-109, -42) | 0.000 | 5,576 | 3480 | -60 (-74, -45) | 0.000 |
| Escitalopram exposed | 320 | 3,324 | -74 (-136, -12) | 0.018 | 6,294 | 3492.38 | -32.87 (-47, -19) | 0.000 | 6,651 | 3501 | -38 (-51, -25) | 0.000 |
| Venlafaxine exposed | 408 | 3,323 | -74 (-130, -18) | 0.009 | 1,218 | 3473.52 | -51.73 (-84, -19) | 0.002 | 3,516 | 3481 | -58 (-77, -39) | 0.000 |
| Mirtazapine exposed | 314 | 3,180 | -217 (-288, -147) | 0.000 | 473 | 3486.30 | -38.95 (-89, 12) | 0.130 | 1,420 | 3480 | -59 (-90, -28) | 0.000 |
| Amitriptyline exposed | 1,262 | 3,359 | -38 (-70, -7) | 0.018 | 1,069 | 3466.04 | -59.21 (-94, -24) | 0.001 | 1,396 | 3495 | -45 (-73, -17) | 0.002 |
| Paroxetine exposed | 726 | 3,333 | -64 (-109, -20) | 0.005 | 250 | 3490.14 | -35.11 (-108, 38) | 0.346 | 1,054 | 3506 | -33 (-67, 0) | 0.049 |
| Duloxetine exposed | 56 | 3,283 | -115 (-283, 54) | 0.181 | 101 | 3442.22 | -83.02 (-209, 43) | 0.197 | 1,096 | 3466 | -74 (-108, -40) | 0.000 |
| Other exposed^[9]^ | 804 | 3,333 | -65 (-106, -24) | 0.002 | 748 | 3527.01 | 1.77 (-38, 41) | 0.930 | 1,121 | 3522 | -18 (-51, 15) | 0.295 |
| Polypharmacy exposed^[10]^ | 1,931 | 3,278 | -120 (-147, -93) | 0.000 | 1,357 | 3460.34 | -64.90 (-95, -35) | 0.000 | 7,984 | 3466 | -73 (-85, -61) | 0.000 |

Figure S6 Individual and pooled odds ratios from fixed effect meta-analyses of individual antidepressants during pregnancy and SGA^[11]^ in the UK, Norway and Sweden for each outcome.

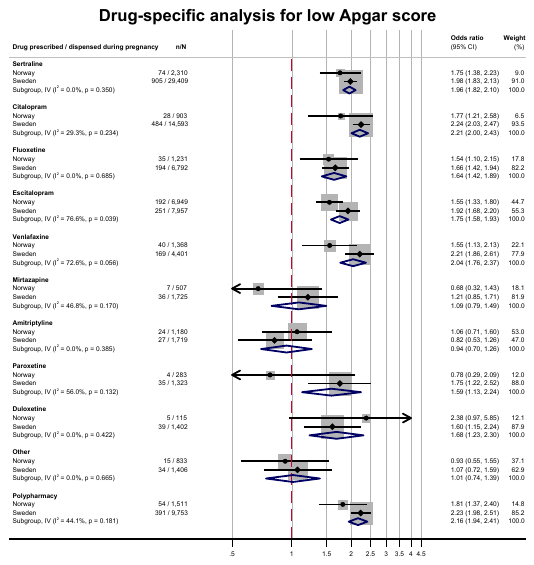

#### 1.7 Discordant sibling analysis

Figure S7 Fixed-effect meta-analysis of estimates generated from sibling models^[12]^ (any antidepressant use during pregnancy compared with discordant siblings) for each outcome in each outcome where available.

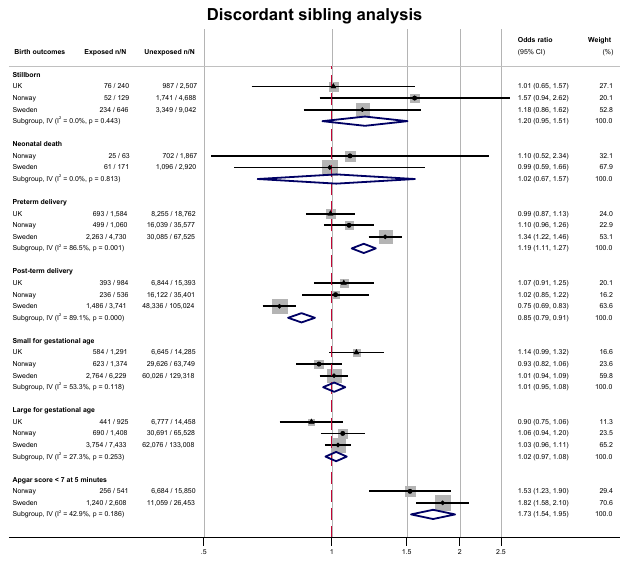

Table S8 Patterns of discordance among siblings in each country for each outcome.

| **Outcome pattern** | **Exposure pattern** | **Country** | **Stillbirth** | **Neonatal death** | **Preterm delivery** | **Post-term delivery** | **SGA** | **LGA** | **Apgar score** |
| --- | --- | --- | --- | --- | --- | --- | --- | --- | --- |
| Concordant, all affected | Concordant, all unexposed | UK | 16 | - | 1,556 | 859 | 3,718 | 4,551 | - |
|  |  | Norway | 8 | 15 | 3,426 | 2,247 | 9,774 | 11,837 | 167 |
|  |  | Sweden | 25 | 6 | 5,754 | 9,474 | 20,009 | 25,302 | 294 |
| Concordant, all affected | Concordant, all exposed | UK | <5 | - | 67 | 20 | 146 | 74 | - |
|  |  | Norway | <5 | <5 | 52 | 13 | 70 | 101 | 8 |
|  |  | Sweden | <5 | <5 | 326 | 68 | 470 | 740 | 56 |
| Concordant, all affected | Discordant | UK | <5 | - | 218 | 91 | 474 | 447 | - |
|  |  | Norway | <5 | <5 | 139 | 37 | 278 | 382 | <5 |
|  |  | Sweden | <5 | <5 | 493 | 290 | 975 | 1,688 | 23 |
| Concordant, all unaffected | Concordant, all unexposed | UK | 144,845 | - | 128,206 | 122,522 | 79,127 | 78,886 | - |
|  |  | Norway | 419,363 | 420,412 | 385,812 | 367,713 | 309,561 | 308,924 | 408,013 |
|  |  | Sweden | 833,452 | 839,249 | 772,006 | 696,106 | 611,384 | 610,470 | 811,817 |
| Concordant, all unaffected | Concordant, all exposed | UK | 2,761 | - | 2,328 | 2,350 | 1,543 | 1,606 | - |
|  |  | Norway | 3,139 | 3,144 | 2,750 | 2,795 | 2,392 | 2,396 | 2,944 |
|  |  | Sweden | 19,600 | 19,842 | 17,418 | 17,263 | 14,622 | 14,361 | 18,238 |
| Concordant, all unaffected | Discordant | UK | 16,111 | - | 13,571 | 13,192 | 8,505 | 8,858 | - |
|  |  | Norway | 13,196 | 13,221 | 11,707 | 11,626 | 9,565 | 9,517 | 12,686 |
|  |  | Sweden | 53,914 | 54,493 | 48,429 | 45,794 | 39,227 | 38,548 | 51,811 |
| Discordant | Concordant, all unexposed | UK | 2,296 | - | 17,395 | 14,485 | 13,062 | 13,608 | - |
|  |  | Norway | 4,571 | 1,804 | 34,704 | 34,897 | 62,551 | 64,271 | 15,460 |
|  |  | Sweden | 8,538 | 2,760 | 64,255 | 101,850 | 124,668 | 127,575 | 25,047 |
| Discordant | Concordant, all exposed | UK | 63 | - | 429 | 184 | 320 | 209 | - |
|  |  | Norway | 38 | 18 | 375 | 139 | 417 | 403 | 224 |
|  |  | Sweden | 298 | 58 | 2,156 | 1,211 | 2,553 | 3,072 | 1,459 |
| Discordant | Discordant | UK | 419 | - | 2,743 | 1,918 | 2,287 | 1,673 | - |
|  |  | Norway | 208 | 108 | 1,558 | 901 | 2,155 | 2,262 | 707 |
|  |  | Sweden | 852 | 273 | 5,844 | 5,704 | 8,326 | 9,794 | 2,555 |

#### Paternal negative control analysis

Figure S8 Fixed-effect meta-analysis of estimates generated from paternal models^[13]^ (any paternal antidepressant use during pregnancy compared with no use) for each outcome in each outcome where available.

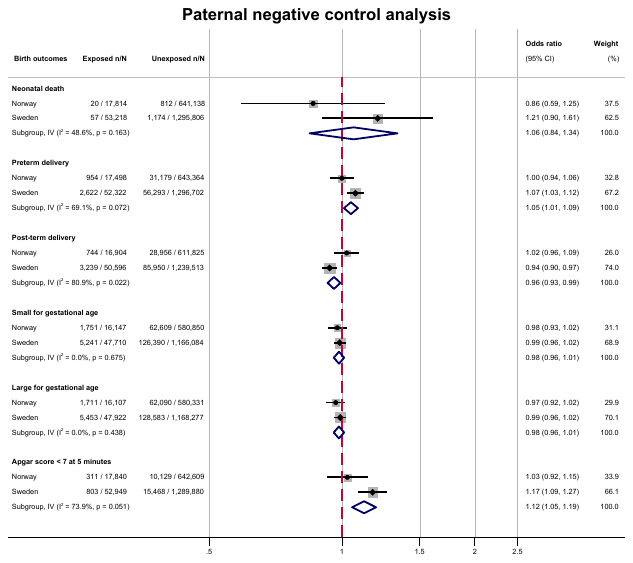

**Table S9** Maternal and paternal characteristics by paternal exposure status.

| **Characteristics** | **Norway** | | **Sweden** | |
| --- | --- | --- | --- | --- |
|  | **Paternal exposed** | **Paternal unexposed** | **Paternal exposed** | **Paternal unexposed** |
| **Total** | 17,958 (100.0) | 646,791 (100.0) | 58,122 (100.0) | 1,404,271 (100.0) |
| **Year of birth** |  |  |  |  |
| 1996-1999 | - | - | - | - |
| 2000-2004 | - | - | - | - |
| 2005-2009 | 1,575 (8.8) | 57,395 (8.9) | 9,727 (16.7) | 271,163 (19.3) |
| 2010-2015 | 9,238 (51.4) | 332,266 (51.4) | 24,444 (42.1) | 618,352 (44.0) |
| 2016-2020 | 7,145 (39.8) | 257,130 (39.8) | 23,951 (41.2) | 514,756 (36.7) |
| **Maternal age at delivery** |  |  |  |  |
| <20 | 297 (1.7) | 8,369 (1.3) | 518 (0.9) | 11,073 (0.8) |
| 20-24 | 2,281 (12.7) | 78,340 (12.1) | 5,357 (9.2) | 145,480 (10.4) |
| 25-29 | 5,218 (29.1) | 207,851 (32.1) | 15,285 (26.3) | 407,537 (29.0) |
| 30-34 | 5,871 (32.7) | 223,960 (34.6) | 20,153 (34.7) | 497,563 (35.4) |
| 35-39 | 3,438 (19.1) | 106,832 (16.5) | 12,990 (22.3) | 273,646 (19.5) |
| 40-44 | 791 (4.4) | 20,386 (3.2) | 3,539 (6.1) | 64,111 (4.6) |
| 45+ | 62 (0.3) | 1,053 (0.2) | 231 (0.4) | 3,970 (0.3) |
| **Maternal educational attainment** |  |  |  |  |
| Compulsory or less | 4,524 (25.2) | 106,748 (16.5) | 23,600 (40.6) | 573,533 (40.8) |
| Secondary | 4,881 (27.2) | 158,156 (24.5) | 20,757 (35.7) | 546,140 (38.9) |
| Post-secondary | 5,706 (31.8) | 243,763 (37.7) | 7,438 (12.8) | 145,479 (10.4) |
| Post-graduate | 2,098 (11.7) | 109,651 (17.0) | 5,387 (9.3) | 108,025 (7.7) |
| Missing (likely educated overseas) | 749 (4.2) | 28,473 (4.4) | 940 (1.6) | 31,094 (2.2) |
| **Household income quintile** |  |  |  |  |
| 1 | - | - | 11,848 (20.4) | 233,330 (16.6) |
| 2 | - | - | 13,454 (23.1) | 271,286 (19.3) |
| 3 | - | - | 11,863 (20.4) | 289,599 (20.6) |
| 4 | - | - | 10,616 (18.3) | 299,349 (21.3) |
| 5 | - | - | 10,338 (17.8) | 310,365 (22.1) |
| Missing | - | - | 3 (0.0) | 342 (0.0) |
| **Maternal country of birth** |  |  |  |  |
| Non-native-born | 4,884 (27.2) | 170,152 (26.3) | 12,726 (21.9) | 365,359 (26.0) |
| Missing | 100 (0.6) | 3,787 (0.6) | 52 (0.1) | 1,056 (0.1) |
| **Maternal BMI** |  |  |  |  |
| Underweight (<18.5 kg/m^2^) | 476 (2.7) | 17,633 (2.7) | 1,363 (2.3) | 32,765 (2.3) |
| Healthy weight (18.5-24.9 kg/m^2^) | 6,895 (38.4) | 273,252 (42.2) | 29,990 (51.6) | 773,049 (55.0) |
| Overweight (25.0-29.9 kg/m^2^) | 3,099 (17.3) | 98,845 (15.3) | 14,295 (24.6) | 335,624 (23.9) |
| Obese (>=30.0 kg/m^2^) | 2,041 (11.4) | 54,395 (8.4) | 8,765 (15.1) | 176,454 (12.6) |
| Missing | 5,447 (30.3) | 202,666 (31.3) | 3,709 (6.4) | 86,379 (6.2) |
| **Maternal smoking during pregnancy** |  |  |  |  |
| Smoker | 1,990 (11.1) | 38,760 (6.0) | 1,250 (2.2) | 14,641 (1.0) |
| Missing | 2,038 (11.3) | 74,858 (11.6) | 3,087 (5.3) | 68,127 (4.9) |
| **Maternal history of stillbirth at the start of pregnancy** |  |  |  |  |
| History of stillbirth | 177 (1.0) | 4,591 (0.7) | 283 (0.5) | 4,907 (0.3) |
| **Parity** |  |  |  |  |
| 0 | 7,029 (39.1) | 272,937 (42.2) | 24,053 (41.4) | 610,217 (43.5) |
| 1 | 6,382 (35.5) | 239,961 (37.1) | 20,587 (35.4) | 521,885 (37.2) |
| 2 | 3,027 (16.9) | 96,411 (14.9) | 8,825 (15.2) | 189,376 (13.5) |
| 3 | 981 (5.5) | 25,191 (3.9) | 2,865 (4.9) | 52,016 (3.7) |
| 4+ | 539 (3.0) | 12,291 (1.9) | 781 (1.3) | 13,453 (1.0) |
| Missing | 0 (0) | 0 (0) | 1,011 (1.7) | 17,324 (1.2) |
| **Maternal indication ever before the start of pregnancy** |  |  |  |  |
| Depression | 5,953 (33.1) | 130,081 (20.1) | 7,135 (12.3) | 78,548 (5.6) |
| Anxiety | 4,307 (24.0) | 94,832 (14.7) | 9,396 (16.2) | 113,784 (8.1) |
| **Medication use in the 12 months before pregnancy** |  |  |  |  |
| Antipsychotics | 398 (2.2) | 6,672 (1.0) | 680 (1.2) | 5,994 (0.4) |
| Anti-seizure medications | 269 (1.5) | 3,776 (0.6) | 666 (1.1) | 7,057 (0.5) |
| **Number of primary care consultations in the 12 months before pregnancy** |  |  |  |  |
| 0 | 1,760 (9.8) | 92,293 (14.3) | - | - |
| 1-3 | 4,673 (26.0) | 203,986 (31.5) | - | - |
| 4-10 | 6,916 (38.5) | 240,525 (37.2) | - | - |
| >10 | 4,609 (25.7) | 109,987 (17.0) | - | - |
| **Induction of labour** |  |  |  |  |
| Induced | 5,471 (30.5) | 178,452 (27.6) | 47,868 (82.4) | 1,183,435 (84.3) |
| Missing | 0 (0) | 0 (0) | 0 (0.0) | 0 (0.0) |
| **Paternal characteristics** |  |  |  |  |
| **Paternal educational attainment** |  |  |  |  |
| Compulsory or less | 5,688 (31.7) | 118,168 (18.3) | 24,405 (42.0) | 647,819 (46.1) |
| Secondary | 6,657 (37.1) | 252,442 (39.0) | 13,841 (23.8) | 374,828 (26.7) |
| Post-secondary | 3,401 (18.9) | 151,577 (23.4) | 10,220 (17.6) | 170,147 (12.1) |
| Postgraduate | 1,798 (10.0) | 100,627 (15.6) | 8,893 (15.3) | 177,400 (12.6) |
| Missing (likely educated overseas) | 414 (2.3) | 23,977 (3.7) | 763 (1.3) | 34,077 (2.4) |
| **Paternal indication ever before the start of pregnancy** |  |  |  |  |
| Depression | 11,149 (62.1) | 50,882 (7.9) | 14,260 (24.5) | 26,253 (1.9) |
| Anxiety | 7,658 (42.6) | 29,612 (4.6) | 18,915 (32.5) | 45,372 (3.2) |

^1^ ICD-10 and ICPC-2 codes in Norway, and ICD-9 and ICD-10 codes in Sweden

Figure S9 Fixed effects meta-analysis of mutually adjusted^[14]^ paternal models in Norway and Sweden where available.

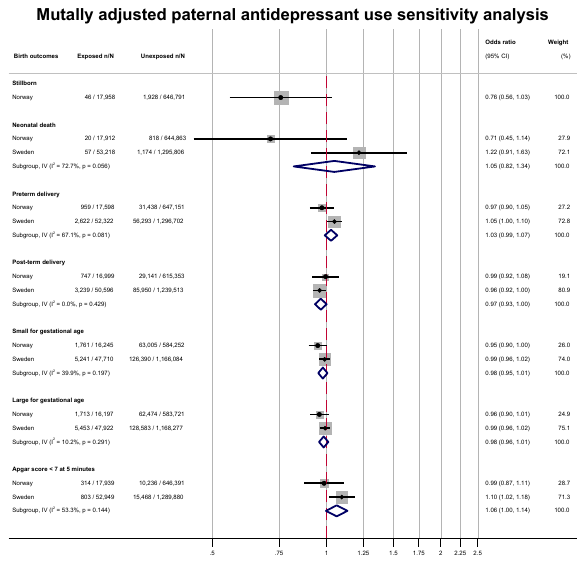

#### Sensitivity analyses

Figure S10 Meta-analysis of each country for additional adjustment^[15]^ sensitivity analyses.

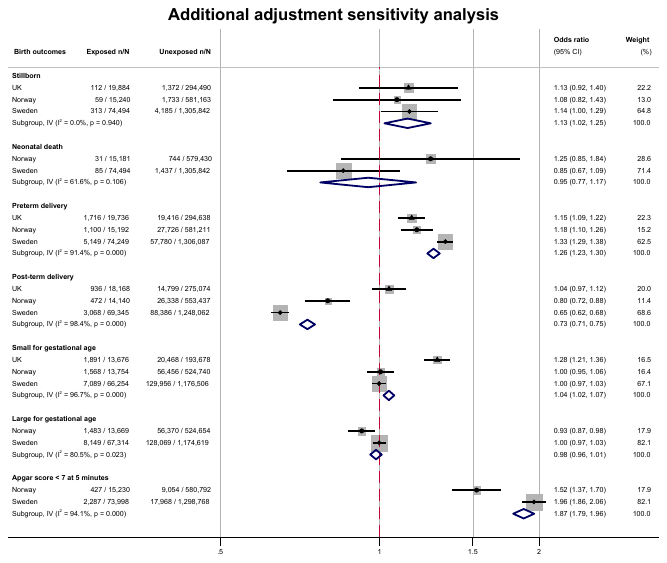

Figure S11 Meta-analysis of adjusted^[16]^ estimates from each country for indication-based sample sensitivity analyses.

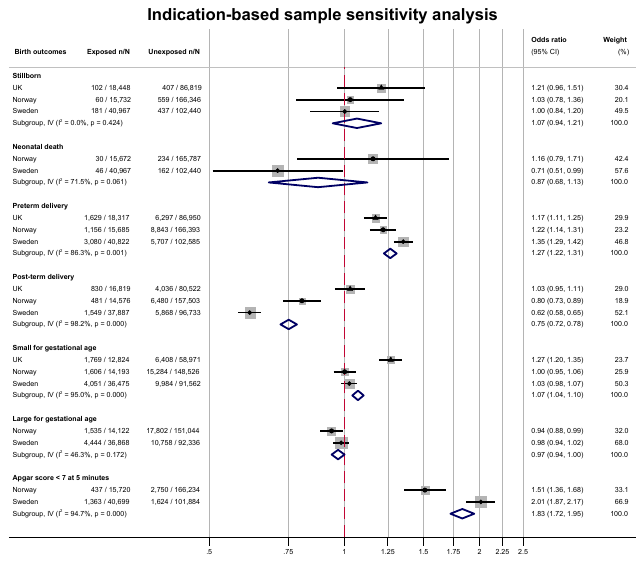

Figure S12 Meta-analysis of adjusted^[17]^ estimates for each country for stratified preterm delivery analyses: moderate-to-late preterm (32-37 weeks’ gestation), very preterm (28-32 weeks’ gestation), and extremely preterm delivery (<28 weeks’ delivery) ^[18]^.

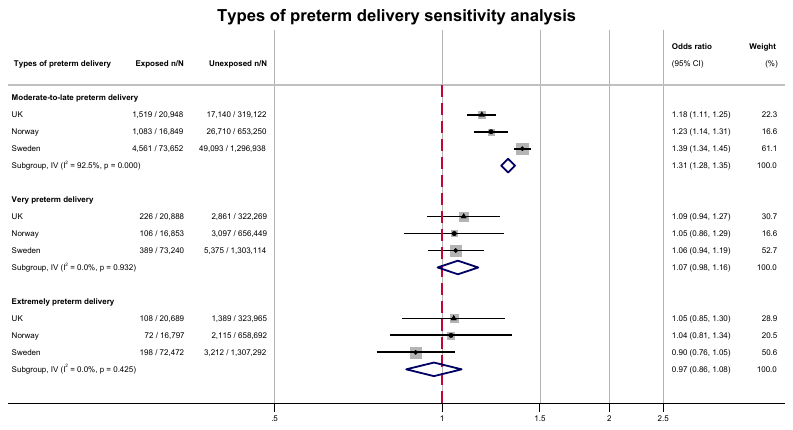

Figure S13 Meta-analysis of adjusted^[19]^ estimates for each country for type of labour initiation among preterm deliveries.

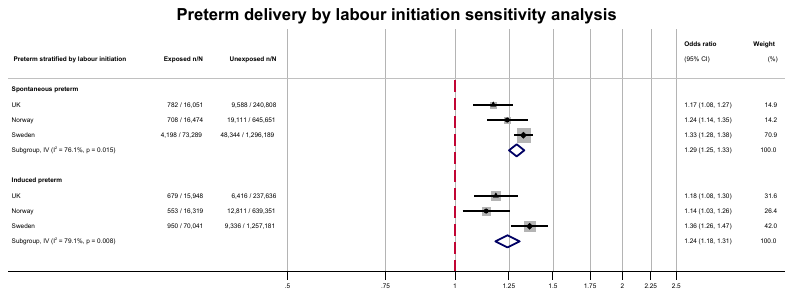

Figure S14 Meta-analysis of adjusted^[20]^ estimates for each country for post-term delivery sensitivity analysis^[21]^ among spontaneous deliveries.

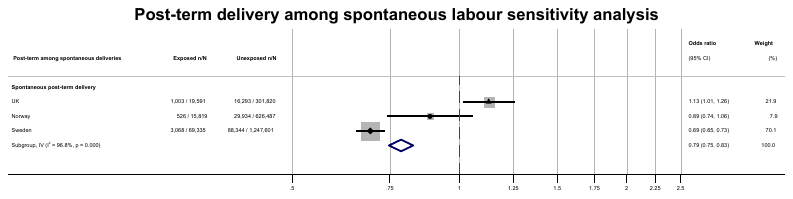

Table S10 SGA and LGA analyses in UK, Norway, and Sweden using different thresholds: from the INTERGROWTH-21 standard and the new Swedish standard.

| **Outcome** | **Country** | **Total^[22]^** | **Exposed n/N (%)** | **Unexposed n/N (%)** | **OR** | **aOR^[23]^** |
| --- | --- | --- | --- | --- | --- | --- |
| SGA using INTERGROWTH-21 | UK | 285,522 | 1,630/ 18,035 (9.04) | 17,666/267,487 (6.60) | 1.41 (1.33-1.48) | 1.24 (1.17-1.32) |
|  | Norway | 515,101 | 672/ 13,021 (5.16) | 23,217/502,080 (4.62) | 1.12 (1.04-1.22) | 1.04 (0.96-1.13) |
|  | Sweden | 1,026,530 | 2,623/54,434 (4.82) | 46,373/972,096 (4.77) | 1.01 (0.97-1.05) | 1.00 (0.95-1.04) |
| SGA using new Swedish standard | UK | 320,453 | 4,676/ 20,024 (23.35) | 59,826/300,429 (19.91) | 1.23 (1.18-1.27) | 1.11 (1.07-1.16) |
|  | Norway | 645,233 | 3,630/ 16,262 (22.32) | 132,846/628,971 (21.12) | 1.07 (1.03-1.12) | 1.05 (1.01-1.10) |
|  | Sweden | 1,259,617 | 11,384/66,190 (17.20) | 207,360/1,193,427 (17.38) | 0.99 (0.97-1.01) | 1.02 (0.99-1.04) |
| LGA using INTERGROWTH-21 | UK | 325,358 | 3,409/ 19,814 (17.21) | 55,723/305,544 (18.24) | 0.93 (0.90-0.97) | 0.93 (0.89-0.97) |
|  | Norway | 651,600 | 4,059/ 16,408 (24.74) | 156,329/635,192 (24.61) | 1.01 (0.97-1.04) | 0.96 (0.92-1.00) |
|  | Sweden | 1,310,350 | 18,525/70,336 (26.34) | 314,291/1,240,014 (25.35) | 1.05 (1.03-1.07) | 0.96 (0.95-0.98) |
| LGA using new Swedish standard | UK | 280,152 | 1,420/ 16,768 (8.47) | 22,781/263,384 (8.65) | 0.98 (0.92-1.03) | 0.99 (0.93-1.05) |
|  | Norway | 539,013 | 818/ 13,450 (6.08) | 29,438/525,563 (5.60) | 1.09 (1.01-1.17) | 0.89 (0.82-0.96) |
|  | Sweden | 1,140,602 | 6,769/61,575 (10.99) | 92,960/1,079,027 (8.62) | 1.31 (1.27-1.35) | 1.12 (1.08-1.15) |

##### UK data management sensitivity analyses

###### Full Pregnancy Register

Table S11 Primary analysis run in the full sample (eligible Pregnancy Register) for variables that are available in the Pregnancy Register in the UK’s CPRD GOLD: stillbirth and gestational age at delivery.

| **Outcome** | **Total complete mums** | **Maternal exposed n/N (%)** | **Maternal unexposed n/N (%)** | **OR** | **aOR^[24]^** |
| --- | --- | --- | --- | --- | --- |
| Stillbirth | 496,496 | 185/ 34,385 (0.54) | 1,991/462,111 (0.43) | 1.25 (1.07-1.45) | 1.21 (1.02-1.43) |
| Neonatal death | - | - | - | - | - |
| Preterm birth^[25]^ | 496,496 | 2,691/ 34,152 (7.88) | 27,957/462,344 (6.05) | 1.33 (1.27-1.39) | 1.11 (1.06-1.17) |
| Post-term birth | 465,848 | 1,385/ 31,694 (4.37) | 21,125/434,154 (4.87) | 0.89 (0.84-0.94) | 0.97 (0.91-1.03) |
| SGA | - | - | - | - | - |
| LGA | - | - | - | - | - |
| Apgar <7 | - | - | - | - | - |

###### Pre- and post-term birth from Pregnancy Register

Table S12 Pre- and post-term analyses among those flagged as pre- or post-term in the full eligible Pregnancy Register sample.

| **Outcome** | **Total complete mums** | **Maternal exposed n/N (%)** | **Maternal unexposed n/N (%)** | **OR** | **aOR^[26]^** |
| --- | --- | --- | --- | --- | --- |
| Preterm birth^[27]^ | 479,559 | 1,352/ 33,185 (4.07) | 12,310/446,374 (2.76) | 1.50 (1.41-1.59) | 1.14 (1.07-1.22) |
| Post-term birth | 479,559 | 1,735/ 33,185 (5.23) | 34,028/446,374 (7.62) | 0.67 (0.64-0.70) | 0.71 (0.67-0.75) |

###### Post-term ≥41 weeks’ gestation

Table S13 Post-term birth defined as ≥41 weeks’ gestation at delivery.

| **Outcome** | **Country** | **Total complete mums** | **Maternal exposed n/N (%)** | **Maternal unexposed n/N (%)** | **OR** | **aOR^[28]^** |
| --- | --- | --- | --- | --- | --- | --- |
| Post-term birth | UK | 321,411 | 3,196/ 19,591 (16.31) | 60,558/301,820 (20.06) | 0.78 (0.75-0.81) | 0.88 (0.84-0.92) |
|  | Norway | 642,306 | 3,276/ 15,819 (20.71) | 167,995/626,487 (26.82) | 0.71 (0.69-0.74) | 0.82 (0.79-0.85) |
|  | Sweden | 1,316,936 | 12,976/ 69,335 (18.71) | 336,205/1,247,601 (26.95) | 0.62 (0.61-0.64) | 0.66 (0.65-0.67) |

1. Maternal models adjusted for year of birth, maternal age, practice-level IMD quintile (UK), maternal ethnicity (UK), maternal educational attainment (Norway and Sweden), household disposable income at the start of pregnancy (Sweden only), smoking during pregnancy (UK and Sweden), maternal BMI at the start of pregnancy (Sweden), previous stillbirth, parity, antipsychotic and anti-seizure medication use in the 12 months before pregnancy, number of primary care consultations in the 12 months before pregnancy (UK and Norway only), depression ever before the start of pregnancy, anxiety ever before the start of pregnancy
2. Trimester-specific models adjusted for year of birth, maternal age, practice-level IMD quintile (UK), maternal ethnicity (UK), maternal educational attainment (Norway and Sweden), household disposable income at the start of pregnancy (Sweden only), smoking during pregnancy (UK and Sweden), maternal BMI at the start of pregnancy (Sweden), previous stillbirth, parity, antipsychotic and anti-seizure medication use in the 12 months before pregnancy, number of primary care consultations in the 12 months before pregnancy (UK and Norway only), depression ever before the start of pregnancy, anxiety ever before the start of pregnancy
3. Models adjusted for year of birth, maternal age, practice-level IMD quintile (UK), maternal ethnicity (UK), maternal educational attainment (Norway and Sweden), household disposable income at the start of pregnancy (Sweden only), smoking during pregnancy (UK and Sweden), maternal BMI at the start of pregnancy (Sweden), previous stillbirth, parity, antipsychotic and anti-seizure medication use in the 12 months before pregnancy, number of primary care consultations in the 12 months before pregnancy (UK and Norway only), depression ever before the start of pregnancy, anxiety ever before the start of pregnancy
4. Adjusted for year of birth, maternal age, smoking during pregnancy (UK and Sweden), maternal ethnicity (UK), previous stillbirth, practice-level IMD quintile, parity, antipsychotic and anti-seizure medication use in the 12 months before pregnancy, number of primary care consultations in the 12 months before pregnancy, depression ever before the start of pregnancy, anxiety ever before the start of pregnancy
5. Drugs included in other: agomelatine, amoxapine, butriptyline, clomipramine, desipramine, dosulepin, doxepin, fluvoxamine, imipramine, iprindole, iproniazide, isocarboxazid, lofepramine, maprotiline, mianserin, moclobemide, nefazodone, nortriptyline, phenelzine, protriptyline, reboxetine, tranylcypromine, trazodone, trimipramine, tryptophan, vortioxetine
6. Polypharmacy defined as patients who filled a prescription for more than one type of antidepressant substance during pregnancy
7. Models adjusted for year of birth, maternal age, practice-level IMD quintile (UK), maternal ethnicity (UK), maternal educational attainment (Norway and Sweden), household disposable income at the start of pregnancy (Sweden only), smoking during pregnancy (UK and Sweden), maternal BMI at the start of pregnancy (Sweden), previous stillbirth, parity, antipsychotic and anti-seizure medication use in the 12 months before pregnancy, number of primary care consultations in the 12 months before pregnancy (UK and Norway only), depression ever before the start of pregnancy, anxiety ever before the start of pregnancy
8. Adjusted for year of birth, maternal age, smoking during pregnancy (UK and Sweden), maternal ethnicity (UK), previous stillbirth, practice-level IMD quintile, parity, antipsychotic and anti-seizure medication use in the 12 months before pregnancy, number of primary care consultations in the 12 months before pregnancy, depression ever before the start of pregnancy, anxiety ever before the start of pregnancy
9. Drugs included in other: agomelatine, amoxapine, butriptyline, clomipramine, desipramine, dosulepin, doxepin, fluvoxamine, imipramine, iprindole, iproniazide, isocarboxazid, lofepramine, maprotiline, mianserin, moclobemide, nefazodone, nortriptyline, phenelzine, protriptyline, reboxetine, tranylcypromine, trazodone, trimipramine, tryptophan, vortioxetine
10. Polypharmacy defined as patients who filled a prescription for more than one type of antidepressant substance during pregnancy
11. Models adjusted for year of birth, maternal age, practice-level IMD quintile (UK), maternal ethnicity (UK), maternal educational attainment (Norway and Sweden), household disposable income at the start of pregnancy (Sweden only), smoking during pregnancy (UK and Sweden), maternal BMI at the start of pregnancy (Sweden), previous stillbirth, parity, antipsychotic and anti-seizure medication use in the 12 months before pregnancy, number of primary care consultations in the 12 months before pregnancy (UK and Norway only), depression ever before the start of pregnancy, anxiety ever before the start of pregnancy
12. Sibling models adjusted for birth year, depression, anxiety, primary healthcare utilisation in the 12 months before pregnancy (UK and Norway), antipsychotic and anti-seizure medication in the 12 months prior to pregnancy (family average minus each siblings value to account for non-shared confounding between siblings), maternal age, maternal educational attainment (Norway and Sweden), house disposable income at the start of pregnancy (Sweden), parity, smoking around the start of pregnancy (UK and Sweden)
13. Paternal models adjusted for year of birth, maternal age, practice-level IMD quintile (UK), maternal ethnicity (UK), maternal and paternal educational attainment (Norway and Sweden), household disposable income at the start of pregnancy (Sweden only), smoking during pregnancy (UK and Sweden), maternal BMI at the start of pregnancy (Sweden), previous stillbirth, parity, antipsychotic and anti-seizure medication use in the 12 months before pregnancy, number of primary care consultations in the 12 months before pregnancy (UK and Norway only), maternal and paternal anxiety ever before the start of pregnancy
14. Paternal models adjusted for year of birth, maternal age, practice-level IMD quintile (UK), maternal ethnicity (UK), maternal and paternal educational attainment (Norway and Sweden), household disposable income at the start of pregnancy (Sweden only), smoking during pregnancy (Sweden), maternal BMI at the start of pregnancy (Sweden), previous stillbirth, parity, antipsychotic and anti-seizure medication use in the 12 months before pregnancy, number of primary care consultations in the 12 months before pregnancy (UK and Norway only), maternal and paternal anxiety ever before the start of pregnancy, maternal antidepressant use during pregnancy
15. Maternal models adjusted for year of birth, maternal age, practice-level IMD quintile (UK), maternal ethnicity (UK), maternal educational attainment (Norway and Sweden), household disposable income at the start of pregnancy (Sweden only), smoking during pregnancy, maternal BMI at the start of pregnancy (Sweden and UK), previous stillbirth (UK and Sweden) parity, antipsychotic and anti-seizure medication use in the 12 months before pregnancy, number of primary care consultations in the 12 months before pregnancy (UK and Norway), depression ever before the start of pregnancy, anxiety ever before the start of pregnancy,
16. Maternal models adjusted for year of birth, maternal age, practice-level IMD quintile (UK), maternal ethnicity (UK), maternal educational attainment (Norway and Sweden), household disposable income at the start of pregnancy (Sweden only), smoking around the start of pregnancy (UK and Sweden), maternal BMI at the start of pregnancy (Sweden), previous stillbirth (UK and Sweden) parity, antipsychotic and anti-seizure medication use in the 12 months before pregnancy, number of primary care consultations in the 12 months before pregnancy (UK and Norway only)
17. Maternal models adjusted for year of birth, maternal age, practice-level IMD quintile (UK), maternal ethnicity (UK), maternal educational attainment (Norway and Sweden), household disposable income at the start of pregnancy (Sweden only), smoking around the start of pregnancy (UK and Sweden), maternal BMI at the start of pregnancy (Sweden), previous stillbirth (UK and Sweden) parity, antipsychotic and anti-seizure medication use in the 12 months before pregnancy, number of primary care consultations in the 12 months before pregnancy (UK and Norway only), depression ever before the start of pregnancy, anxiety ever before the start of pregnancy
18. In the moderate-to-late preterm models, those who initiated antidepressants after 36+6 weeks’ gestation were considered unexposed, in the very preterm models those initiated after 31+6 weeks’ gestation were considered unexposed, and in the extremely preterm models those who initiated after 27+6 weeks’ gestation were considered unexposed (due to these “exposed” pregnancies having zero chance to experience the outcome)
19. Maternal models adjusted for year of birth, maternal age, practice-level IMD quintile (UK), maternal ethnicity (UK), maternal educational attainment (Norway and Sweden), household disposable income at the start of pregnancy (Sweden only), smoking around the start of pregnancy (UK and Sweden), maternal BMI at the start of pregnancy (Sweden), previous stillbirth (UK and Sweden) parity, antipsychotic and anti-seizure medication use in the 12 months before pregnancy, number of primary care consultations in the 12 months before pregnancy (UK and Norway only), depression ever before the start of pregnancy, anxiety ever before the start of pregnancy
20. Maternal models adjusted for year of birth, maternal age, practice-level IMD quintile (UK), maternal ethnicity (UK), maternal educational attainment (Norway and Sweden), household disposable income at the start of pregnancy (Sweden only), smoking around the start of pregnancy (UK and Sweden), maternal BMI at the start of pregnancy (Sweden), previous stillbirth (UK and Sweden) parity, antipsychotic and anti-seizure medication use in the 12 months before pregnancy, number of primary care consultations in the 12 months before pregnancy (UK and Norway only), depression ever before the start of pregnancy, anxiety ever before the start of pregnancy
21. Stratified on spontaneous labour and reaching term
22. Excluding pregnancies that occur in the end of the study period (dropped pregnancies whose start date + 42 weeks’ gestation > the latest pregnancy start date in the sample)
23. Adjusted for year of birth, maternal age, practice-level IMD quintile (UK), maternal ethnicity (UK), maternal educational attainment (Norway and Sweden), household disposable income at the start of pregnancy (Sweden only), smoking during pregnancy (UK and Sweden), maternal BMI at the start of pregnancy (Sweden), previous stillbirth, parity, antipsychotic and anti-seizure medication use in the 12 months before pregnancy, number of primary care consultations in the 12 months before pregnancy (UK and Norway only), depression ever before the start of pregnancy, anxiety ever before the start of pregnancy
24. Maternal models adjusted for year of birth, maternal age, smoking during pregnancy, maternal ethnicity, practice-level IMD quintile, parity, antipsychotic and anti-seizure medication use in the 12 months before pregnancy, number of primary care consultations in the 12 months before pregnancy, depression ever before the start of pregnancy, anxiety ever before the start of pregnancy
25. Those newly exposed after 37 weeks’ gestation considered unexposed in these analyses
26. Maternal models adjusted for year of birth, maternal age, practice-level IMD quintile, maternal ethnicity (UK), smoking during pregnancy (UK and Sweden), previous stillbirth, parity, antipsychotic and anti-seizure medication use in the 12 months before pregnancy, number of primary care consultations in the 12 months before pregnancy (UK and Norway), depression ever before the start of pregnancy, anxiety ever before the start of pregnancy
27. Those newly exposed after 37 weeks’ gestation considered unexposed in these analyses
28. Maternal models adjusted for year of birth, maternal age, practice-level IMD quintile, maternal ethnicity, smoking around the start of pregnancy, previous stillbirth, parity, antipsychotic and anti-seizure medication use in the 12 months before pregnancy, number of primary care consultations in the 12 months before pregnancy, depression ever before the start of pregnancy, anxiety ever before the start of pregnancy
